# Age-related changes in physiology in individuals with bipolar disorder

**DOI:** 10.1101/2021.03.05.21252987

**Authors:** Julian Mutz, Cathryn M. Lewis

## Abstract

**Objectives:** Individuals with bipolar disorder have reduced life expectancy and may experience accelerated biological ageing. In individuals with bipolar disorder and healthy controls, we examined differences in age-related changes in physiology.

**Methods:** UK Biobank recruited >500,000 participants, aged 37-73, between 2006–2010. Generalised additive models were used to examine associations between age and grip strength, cardiovascular function, body composition, lung function and bone mineral density. Analyses were conducted separately in males and females with bipolar disorder compared to healthy controls.

**Results:** Analytical samples included up to 272,462 adults (mean age = 56.04 years, SD = 8.15; 49.51% females). We found statistically significant differences between bipolar disorder cases and controls for grip strength, blood pressure, pulse rate and body composition, with standardised mean differences of up to −0.238 (95% CI −0.282 to −0.193). There was limited evidence of differences in lung function, heel bone mineral density or arterial stiffness. Case-control differences were most evident for age-related changes in cardiovascular function (in both sexes) and body composition (in females). These differences did not uniformly narrow or widen with age and differed by sex. For example, the difference in systolic blood pressure between male cases and controls was −1.3 mmHg at age 50 and widened to −4.7 mmHg at age 65. Diastolic blood pressure in female cases was 1.2 mmHg higher at age 40 and −1.2 mmHg lower at age 65.

**Conclusions:** Differences in ageing trajectories between bipolar disorder cases and healthy controls were most evident for cardiovascular and body composition measures and differed by sex.

## Introduction

Individuals with bipolar disorder, on average, die prematurely, with some estimates suggesting between 11 to 20 fewer years of life expectancy from age 15. A study of nationwide registry data from Denmark found that bipolar disorder patients between the age of 25 to 45 had remaining life expectancies that were 12.0 to 8.7 years shorter in males and 10.6 to 8.3 years shorter in females, than in the general population^1^. Data from a secondary mental health care register in the UK suggested that individuals with a diagnosis of bipolar disorder had life expectancies from age 15 that were reduced by 10.1 years in males and 11.2 years in females patients compared to the general population^2^.

The increased mortality observed in individuals with bipolar disorder is partially related to higher rates of suicide^3^, and to a large extent due to higher prevalence of cardiovascular disease and other medical comorbidities^4^. Previous studies suggest that individuals with bipolar disorder have higher rates of cardiovascular disease^5^, osteoporosis^6^, obesity^7^, diabetes^8^ and chronic obstructive pulmonary disease^9^.

Bipolar disorder might also be characterised by accelerated biological ageing, supported, for example, by the observation that the ratio of remaining life expectancy in bipolar disorder and the general population decreases with age, suggesting that individuals with bipolar disorder start losing life-years in their early to mid-adulthood^1^. Moreover, death from natural causes increases substantially from early to mid-adulthood^10^. Several studies have examined proxies for biological ageing, for instance shortened leukocyte telomere length, in bipolar disorder^11,12^. However, less is known about age-related changes in physiological measures in individuals with bipolar disorder. In a previous study we have reported on age-related changes across 15 physiological measures in individuals with lifetime depression compared to healthy controls^13^.

The aim of this study was to examine associations between age and multiple physiological measures, and to test for differences in these measures between middle-aged and older adults with bipolar disorder and healthy controls across age. Given major sex differences in physiology, life expectancy^14^, age-related diseases^14^, and medical comorbidities^15^, we conducted all analyses separately in males and females.

Using data from up to 272,462 participants in the UK Biobank with physiological data collected during the baseline study visit, the following questions were examined for each measure: Are there average differences between males and females with bipolar disorder and healthy controls? Do age-related changes differ between males and females with bipolar disorder and healthy controls?

## Methods

### Study population

The UK Biobank is a prospective study of >500,000 UK residents aged 37–73 at baseline, recruited between 2006–2010. Details of the study rationale and design have been reported elsewhere^16^. Briefly, individuals registered with the UK National Health Service (NHS) and living within a 25-mile (∼40 km) radius of one of 22 assessment centres were invited to participate (9,238,453 postal invitations sent). Participants provided information on their sociodemographic characteristics, lifestyle and medical history. They also underwent physical examination including, for example, height, weight and blood pressure measurement. Linked hospital inpatient records are available for most participants and primary care records are currently available for half of participants. A subset of 157,366 out of 339,092 invited participants (46%) completed an online follow-up mental health questionnaire (MHQ) between 2016-2017, covering 31% of all participants.

### Exposures

Age at baseline assessment was the primary explanatory variable.

Lifetime bipolar disorder was assessed with the MHQ that included several questions on mania and hypomania in combination with the Composite International Diagnostic Interview Short Form (CIDI-SF) to assess lifetime depression (Supplement 1)^17^. We also included in our definition of lifetime bipolar disorder individuals who had self-reported “mania/bipolar disorder/manic depression” during the nurse-led interview at baseline (field 20002) or “mania, hypomania, bipolar or manic-depression” in response to a single-item question on the MHQ (field 20544), participants with a hospital inpatient record containing an ICD-10 code for mania or bipolar disorder (F30-F31; Supplement 2), participants with a primary care record containing a Read v2 or CTV3 code for bipolar disorder (for diagnostic codes and data extraction procedures, see ^18^), and those who were classified as individuals with probable bipolar disorder according to Smith et al.^19^ based on additional questions that were introduced during the later stages of the baseline assessment (Supplement 3). We excluded individuals with any record of psychosis.

Healthy controls were defined as individuals who did not meet our criteria for bipolar disorder and had no record of other mental disorders: (i) had not self-reported “schizophrenia”, “depression”, “anxiety/panic attacks”, “obsessive compulsive disorder”, “anorexia/bulimia/other eating disorder”, “post-traumatic stress disorder” during the nurse-led interview at baseline (field 20002); (ii) reported no mental disorder in response to a single-item question on the MHQ (field 20544); (iii) had self-reported no current psychotropic medication use at baseline (Supplement 4)^20^; (iv) had no linked hospital inpatient record that contained any ICD-10 Chapter V code except organic causes or substance use (F20-F99); (v) had no primary care record containing a diagnostic code for mental disorder^18^; (vi) were not classified as individuals with probable major depressive disorder according to Smith et al.^19^; (vi) no Patient Health Questionnaire-9 (PHQ-9) sum score of ≥5.

### Outcomes

We examined 15 continuous physiological measures obtained at the baseline assessment, including maximal hand-grip strength, systolic and diastolic resting blood pressure, resting pulse rate, body mass index (BMI), waist-hip ratio, fat mass, fat-free mass, body fat percentage, peak expiratory flow, forced vital capacity (FVC), forced expiratory volume in one second (FEV_1_), FVC/FEV_1_ ratio, heel bone mineral density and arterial stiffness. Details on these measures that have previously been reported in our study on age-related changes in physiology in depression^13^ are included in Supplement 5.

Briefly, hand-grip strength in whole kilogram force units was measured using a Jamar J00105 hydraulic hand dynamometer. Seated systolic and diastolic resting blood pressure in millimetres of mercury (mmHg) was measured twice using an Omron 705 IT digital blood pressure monitor. Resting pulse rate in beats per minute was recorded during the blood pressure measurements using the Omron 705 IT device or, exceptionally, a manual sphygmomanometer. Weight and body composition measurements were obtained with a Tanita BC-418 MA body composition analyser or, in limited cases, using a manual scale. Standing height was measured using a Seca 202 height measure. Waist and hip circumference in cm were measured using a Wessex non-stretchable sprung tape. Other body composition measures were derived from these variables. Volumetric measures of lung function were quantified using breath spirometry with a Vitalograph Pneumotrac 6800. Heel bone mineral density was estimated by quantitative ultrasound assessment of the calcaneus using a Sahara Clinical Bone Sonometer. Resting pulse wave velocity, used to derive an index of arterial stiffness, was measured using finger photoplethysmography with a PulseTrace PCA2 infra-red sensor.

### Exclusion criteria

Participants for whom their genetic sex, inferred from genotype information on the Y and X chromosomes, and self-reported sex did not match were excluded. Participants with missing data or who responded “do not know” or “prefer not to answer” to any covariates assessed were also excluded.

### Covariates

Covariates included ethnicity (White, Asian, Black, Chinese, mixed-race or other), gross annual household income (<£18,000, £18,000–30,999, £31,000–51,999, £52,000–100,000 or >£100 000), physical activity (number of days per week spent walking, engaging in moderate-intensity physical activity or engaging in vigorous-intensity physical activity for ≥10 minutes continuously), smoking status (never, former or current), alcohol intake frequency (never, special occasions only, one to three times a month, once or twice a week, three or four times a week or daily or almost daily), sleep duration (hours per day) and, for cardiovascular measures, current use of antihypertensive medications (yes/no; derived from self-reported medication codes) (Supplement 6). A more detailed description of these variables is available in our previous publication^21^.

### Statistical analyses

Analyses were pre-specified prior to inspection of the data (preregistration: osf.io/rjvqt) and algorithms were tested on simulated data. Statistical analyses were conducted using R (version 3.6.0).

Sample characteristics were summarised using means and standard deviations or counts and percentages. We also present the number of individuals who met our criteria for bipolar disorder and healthy control. Density plots of all physiological measures stratified by bipolar disorder case status are presented separately for males and females. Overall differences between the sexes and between cases and controls were estimated using standardised mean differences (± 95% confidence intervals).

We examined the relationship between each physiological measure and age using generalised additive models (GAMs) with the ‘mgcv’ package in R^22^. GAMs are flexible modelling approaches that allow for the relationship between an outcome variable and a continuous exposure to be represented by a non-linear smooth curve while adjusting for covariates. This approach is particularly useful if a linear model does not capture key aspects of the relationship between variables and attempts to achieve maximum goodness-of-fit while maintaining parsimony of the fitted curve to minimize overfitting. Smoothing parameters were selected using the restricted maximum likelihood method and we used the default option of ten basis functions to represent smooth terms in each model. Each measure was modelled against a penalised regression spline function of age with separate smooths for individuals with bipolar disorder and healthy controls.

Two models were fitted for males and females separately:

- Unadjusted model: physiological measure ∼ bipolar disorder + s(age, by bipolar disorder).
- Adjusted model: physiological measure ∼ bipolar disorder + s(age, by bipolar disorder) + covariates (ethnicity, household income, physical activity, smoking status, alcohol intake frequency and sleep duration).

where s(age, by bipolar disorder) represents the smooth function for age, stratified by bipolar disorder status. For cardiovascular measurements, the adjusted model additionally included self-reported antihypertensive medication use.

To formally test whether the relationship between physiological measures and age differed between individuals with bipolar disorder and healthy controls, we also fitted models that included a reference smooth for healthy controls and a difference smooth for individuals with bipolar disorder compared to healthy controls. For this analysis, bipolar disorder status was coded as an ordered factor in R. If the difference smooth differs from zero, the physiological measure follows a different trend with age in individuals with bipolar disorder and healthy controls.

Adjusted *p*-values were calculated using the p.adjust() command in R to account for multiple testing across each set of analyses of the 15 physiological measures. Two methods were used: (1) Bonferroni and (2) Benjamini & Hochberg^23^, all two-tailed with α = .05 and false discovery rate of 5%, respectively.

To address potential concerns about the validity and reliability of our case definition, we conducted sensitivity analyses in which we restricted our sample to (i) individuals with bipolar disorder according to at least two different measures and (ii) individuals with bipolar disorder status assessed using the MHQ. These sensitivity analyses were not preregistered.

## Results

### Study population

Of the 502,521 UK Biobank participants, 392,467 (78.1%) had complete data on all covariates. After excluding individuals who did not meet our inclusion criteria or whose bipolar disorder status could not be determined, the main analytical sample included 272,462 adults. Sample sizes for physiological measures that were collected in subsets of the UK Biobank ranged from 87,463 to 187,387 participants (Figure 1).

**Figure 1.**
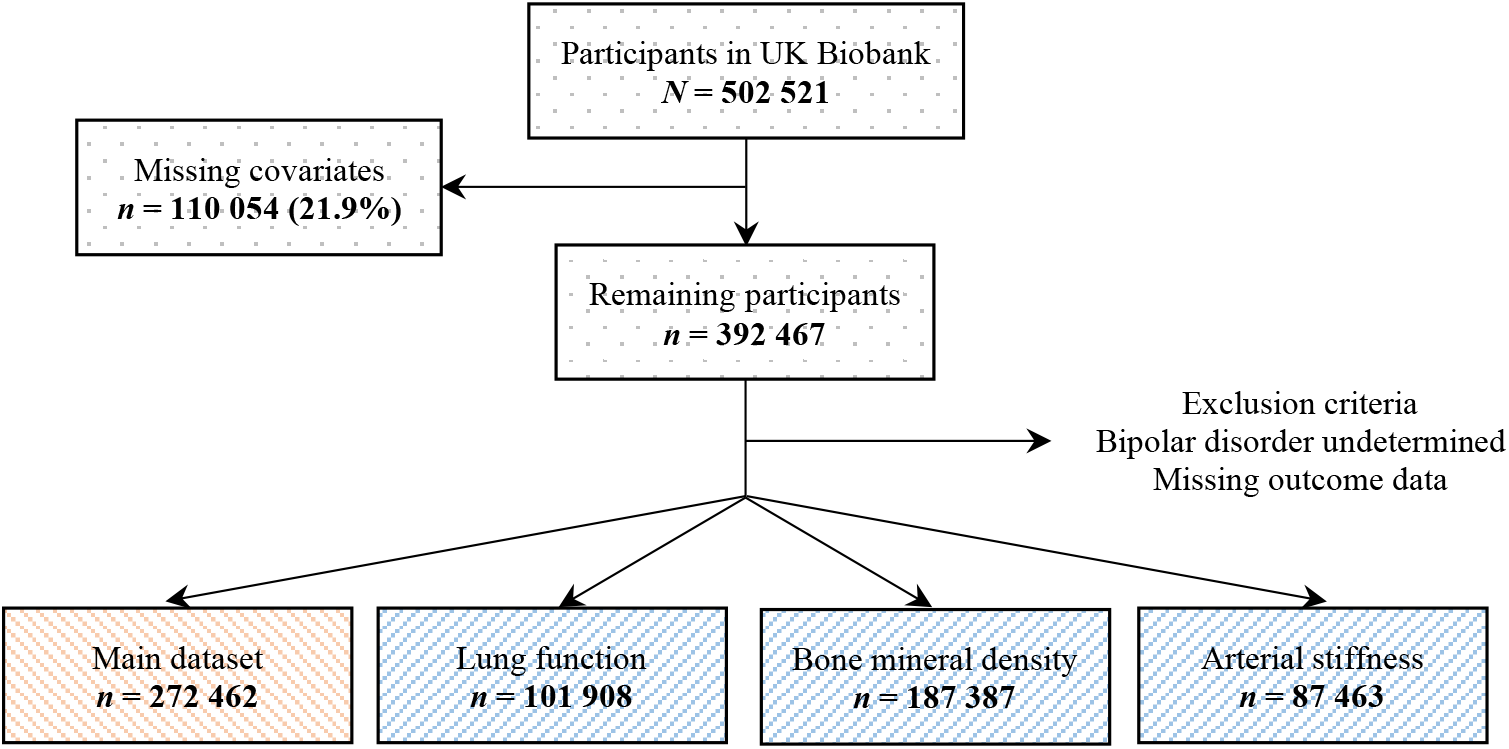
Flowchart of study population. The main dataset included hand-grip strength, blood pressure, pulse rate and measures of body composition.

### Sample characteristics

The average age of participants in our main dataset was 56.04 years (SD = 8.15) and 49.51% of participants were female. About 1.33% of participants (*n* = 3,629) met criteria for bipolar disorder, 54.67% (*n* = 1,984) of whom were female. Case numbers for each data source are presented in Supplement 6. Descriptive statistics and density plots of the physiological measures stratified by sex and bipolar disorder status are presented in Table 1 and Figure 2, respectively. Descriptive statistics of the covariates are presented in the Supplement 7.

**Table 1.**
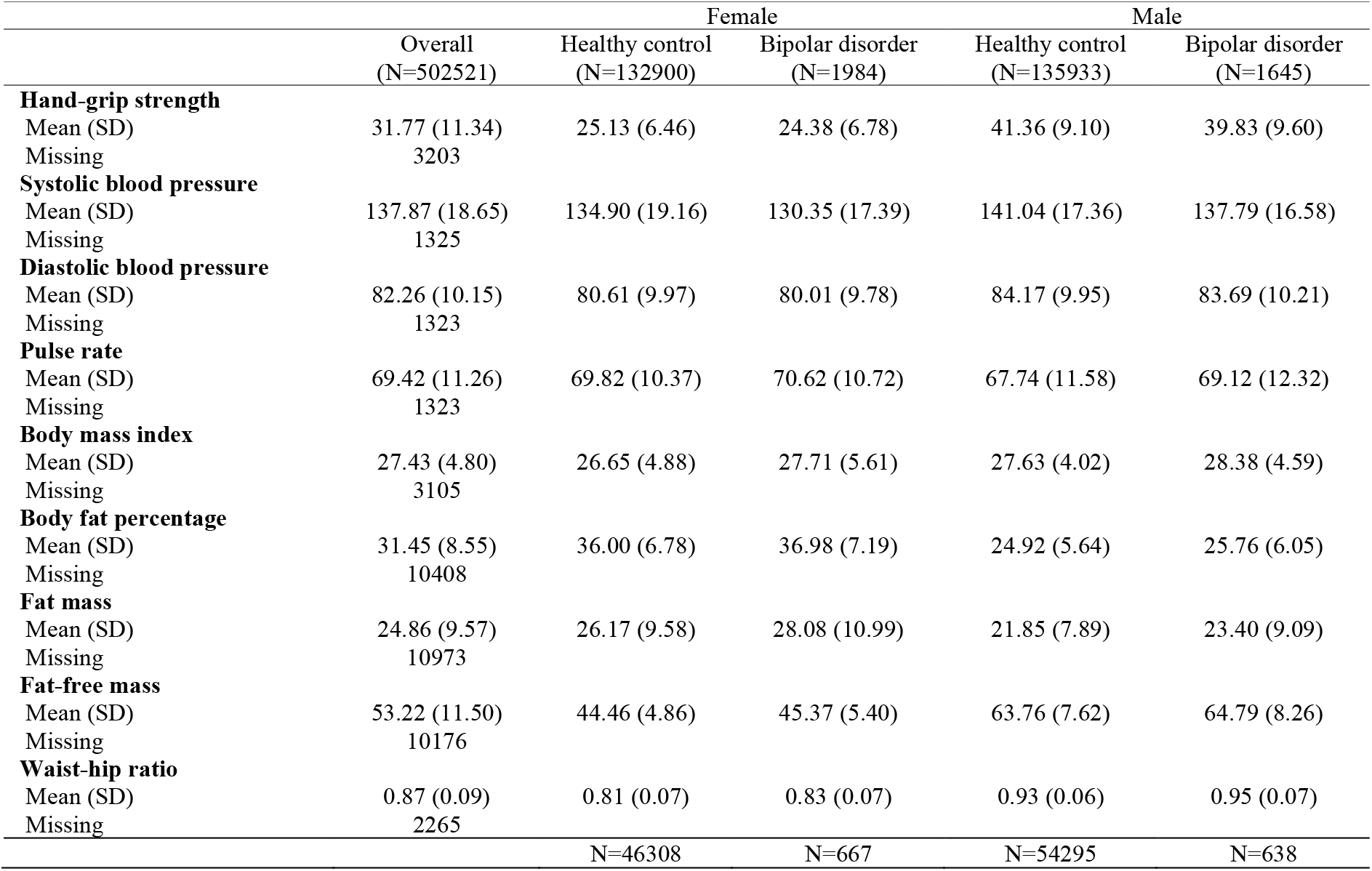
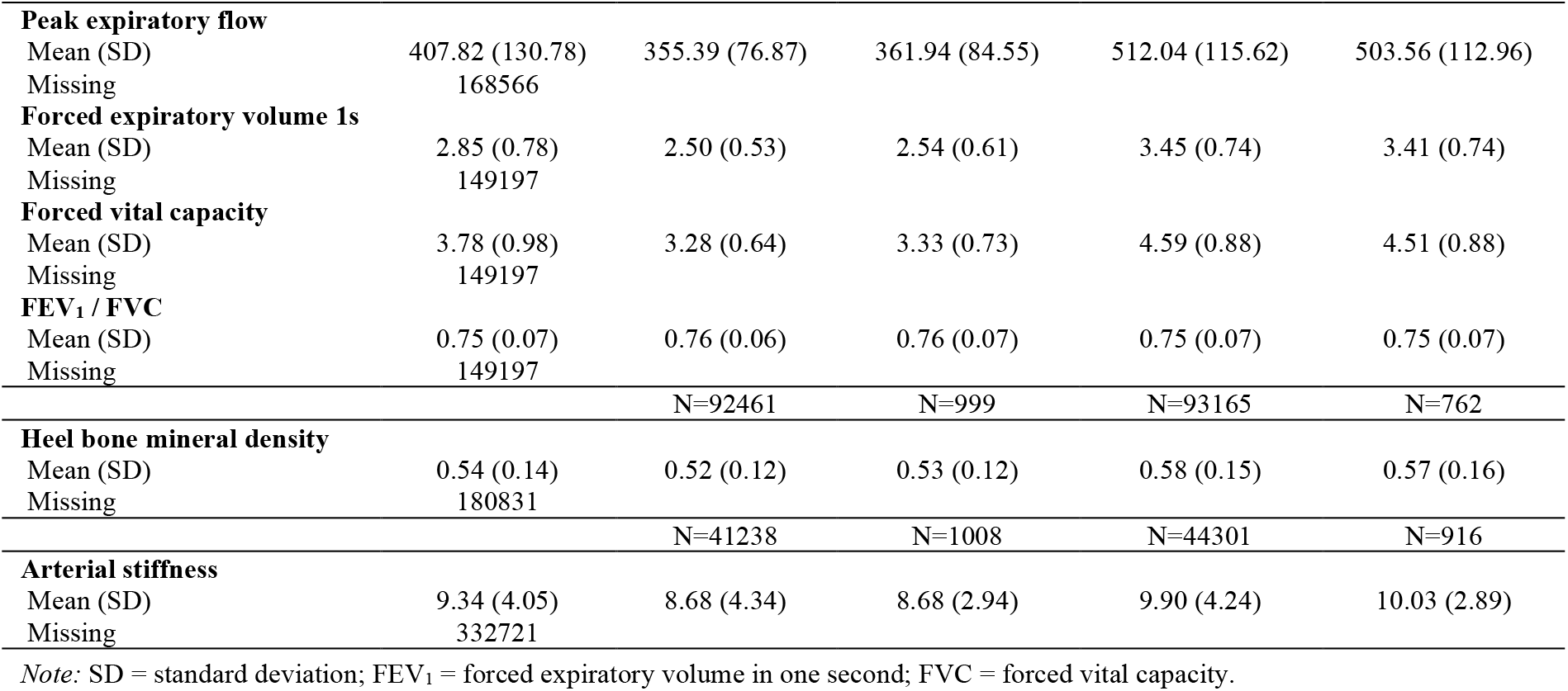
Sample characteristics

|  |  | Female |  | Male |  |
| --- | --- | --- | --- | --- | --- |
|  | Overall<br>(N=502521) | Healthy control<br>(N=132900) | Bipolar disorder<br>(N=1984) | Healthy control<br>(N=135933) | Bipolar disorder<br>(N=1645) |
| **Hand-grip strength** |  |  |  |  |  |
| Mean (SD) | 31.77 (11.34) | 25.13 (6.46) | 24.38 (6.78) | 41.36 (9.10) | 39.83 (9.60) |
| Missing | 3203 |  |  |  |  |
| **Systolic blood pressure** |  |  |  |  |  |
| Mean (SD) | 137.87 (18.65) | 134.90 (19.16) | 130.35 (17.39) | 141.04 (17.36) | 137.79 (16.58) |
| Missing | 1325 |  |  |  |  |
| **Diastolic blood pressure** |  |  |  |  |  |
| Mean (SD) | 82.26 (10.15) | 80.61 (9.97) | 80.01 (9.78) | 84.17 (9.95) | 83.69 (10.21) |
| Missing | 1323 |  |  |  |  |
| **Pulse rate** |  |  |  |  |  |
| Mean (SD) | 69.42 (11.26) | 69.82 (10.37) | 70.62 (10.72) | 67.74 (11.58) | 69.12 (12.32) |
| Missing | 1323 |  |  |  |  |
| **Body mass index** |  |  |  |  |  |
| Mean (SD) | 27.43 (4.80) | 26.65 (4.88) | 27.71 (5.61) | 27.63 (4.02) | 28.38 (4.59) |
| Missing | 3105 |  |  |  |  |
| **Body fat percentage** |  |  |  |  |  |
| Mean (SD) | 31.45 (8.55) | 36.00 (6.78) | 36.98 (7.19) | 24.92 (5.64) | 25.76 (6.05) |
| Missing | 10408 |  |  |  |  |
| **Fat mass** |  |  |  |  |  |
| Mean (SD) | 24.86 (9.57) | 26.17 (9.58) | 28.08 (10.99) | 21.85 (7.89) | 23.40 (9.09) |
| Missing | 10973 |  |  |  |  |
| **Fat-free mass** |  |  |  |  |  |
| Mean (SD) | 53.22 (11.50) | 44.46 (4.86) | 45.37 (5.40) | 63.76 (7.62) | 64.79 (8.26) |
| Missing | 10176 |  |  |  |  |
| **Waist-hip ratio** |  |  |  |  |  |
| Mean (SD) | 0.87 (0.09) | 0.81 (0.07) | 0.83 (0.07) | 0.93 (0.06) | 0.95 (0.07) |
| Missing | 2265 |  |  |  |  |
|  |  | N=46308 | N=667 | N=54295 | N=638 |
| **Peak expiratory flow** |  |  |  |  |  |
| Mean (SD) | 407.82 (130.78) | 355.39 (76.87) | 361.94 (84.55) | 512.04 (115.62) | 503.56 (112.96) |
| Missing | 168566 |  |  |  |  |
| **Forced expiratory volume 1s** |  |  |  |  |  |
| Mean (SD) | 2.85 (0.78) | 2.50 (0.53) | 2.54 (0.61) | 3.45 (0.74) | 3.41 (0.74) |
| Missing | 149197 |  |  |  |  |
| **Forced vital capacity** |  |  |  |  |  |
| Mean (SD) | 3.78 (0.98) | 3.28 (0.64) | 3.33 (0.73) | 4.59 (0.88) | 4.51 (0.88) |
| Missing | 149197 |  |  |  |  |
| **FEV<sub>1</sub> / FVC** |  |  |  |  |  |
| Mean (SD) | 0.75 (0.07) | 0.76 (0.06) | 0.76 (0.07) | 0.75 (0.07) | 0.75 (0.07) |
| Missing | 149197 |  |  |  |  |
|  |  | N=92461 | N=999 | N=93165 | N=762 |
| **Heel bone mineral density** |  |  |  |  |  |
| Mean (SD) | 0.54 (0.14) | 0.52 (0.12) | 0.53 (0.12) | 0.58 (0.15) | 0.57 (0.16) |
| Missing | 180831 |  |  |  |  |
|  |  | N=41238 | N=1008 | N=44301 | N=916 |
| **Arterial stiffness** |  |  |  |  |  |
| Mean (SD) | 9.34 (4.05) | 8.68 (4.34) | 8.68 (2.94) | 9.90 (4.24) | 10.03 (2.89) |
| Missing | 332721 |  |  |  |  |

Note: SD = standard deviation; FEV<sub>1</sub> = forced expiratory volume in one second; FVC = forced vital capacity.

**Figure 2.**
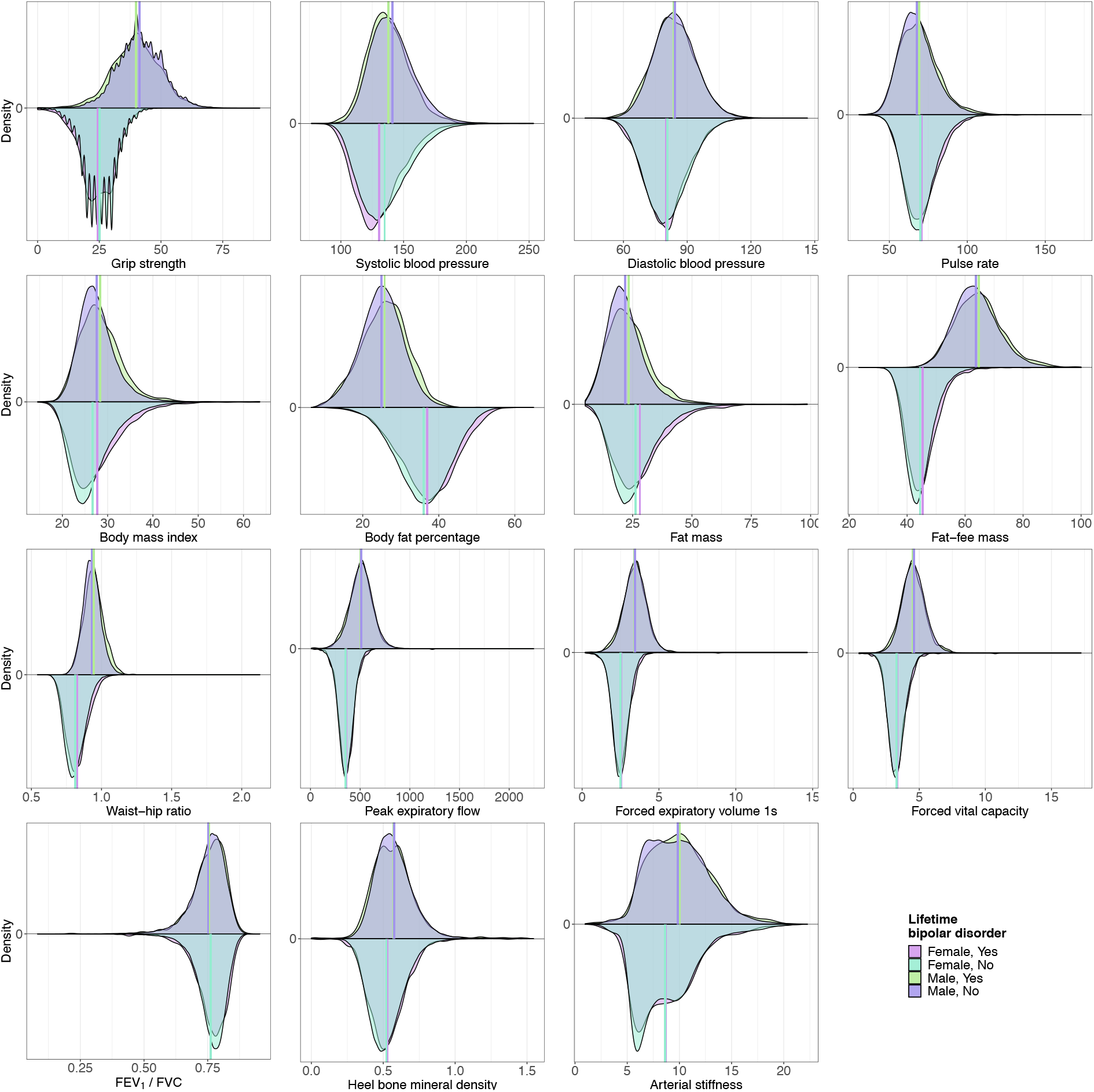
Physiological measures of males and females with bipolar disorder and healthy controls. Arterial stiffness was truncated at the 99.9th %ile for the purpose of this figure due to a small number of very large values (88 observations were outside graph maximum). FEV_1_ = forced expiratory volume in one second; FVC = forced vital capacity.

Sex differences were moderate to large for all physiological measures (all *p*_Bonf._ < 0.001). Females, on average, had lower hand-grip strength, blood pressure, BMI, fat-free mass, waist-hip ratio, peak expiratory flow, FEV_1_, forced vital capacity, heel bone mineral density and arterial stiffness than males. Their pulse rate, body fat percentage, fat mass and FEV_1_/FVC ratio was higher than in males (Table 2).

**Table 2.** Sex differences in physiological measures (females compared to males)

| Variable | SMD | 95% CI |  |
| --- | --- | --- | --- |
| Hand-grip strength | -2.051 | -2.060 | -2.042 |
| Systolic blood pressure | -0.338 | -0.345 | -0.330 |
| Diastolic blood pressure | -0.358 | -0.366 | -0.351 |
| Pulse rate | 0.189 | 0.181 | 0.196 |
| Body mass index | -0.216 | -0.224 | -0.209 |
| Body fat percentage | 1.777 | 1.768 | 1.786 |
| Fat mass | 0.492 | 0.484 | 0.499 |
| Fat-free mass | -3.012 | -3.023 | -3.001 |
| Waist-hip ratio | -1.795 | -1.804 | -1.786 |
| Peak expiratory flow | -1.570 | -1.584 | -1.556 |
| Forced expiratory volume 1s | -1.466 | -1.480 | -1.452 |
| Forced vital capacity | -1.676 | -1.691 | -1.662 |
| FEV<sub>1</sub> / FVC | 0.156 | 0.144 | 0.169 |
| Heel bone mineral density | -0.407 | -0.416 | -0.398 |
| Arterial stiffness | -0.288 | -0.301 | -0.274 |

Note: Analyses included cases plus controls. SMD = standardised mean difference; CI = confidence interval; FEV<sub>1</sub> = forced expiratory volume in one second; FVC = forced vital capacity. All Bonferroni-adjusted $p$ -values for Welch's $t$ -test $< 0.001$ .

We also found statistically significant differences between individuals with bipolar disorder and healthy controls for multiple physiological measures. Female cases, on average, had lower hand-grip strength and lower blood pressure. Their pulse rate, BMI, body fat percentage, fat mass, fat-free mass and waist-hip ratio were higher than in healthy controls. We did not find evidence of differences in lung function, heel bone mineral density or arterial stiffness after correcting for multiple testing. Male cases had lower hand-grip strength, systolic blood pressure and forced vital capacity, while their pulse rate, BMI, body fat percentage, fat mass, fat-free mass and waist-hip ratio were higher than in healthy controls. We did not find evidence that diastolic blood pressure, other measures of lung function, heel bone mineral density or arterial stiffness differed between male cases and controls. The differences observed between cases and controls, although highly statistically significant, were subtle, with the largest effect size corresponding to a standardised mean difference of −0.24 (95% CI −0.28 to − 0.19) for systolic blood pressure in females (Table 3).

**Table 3.**
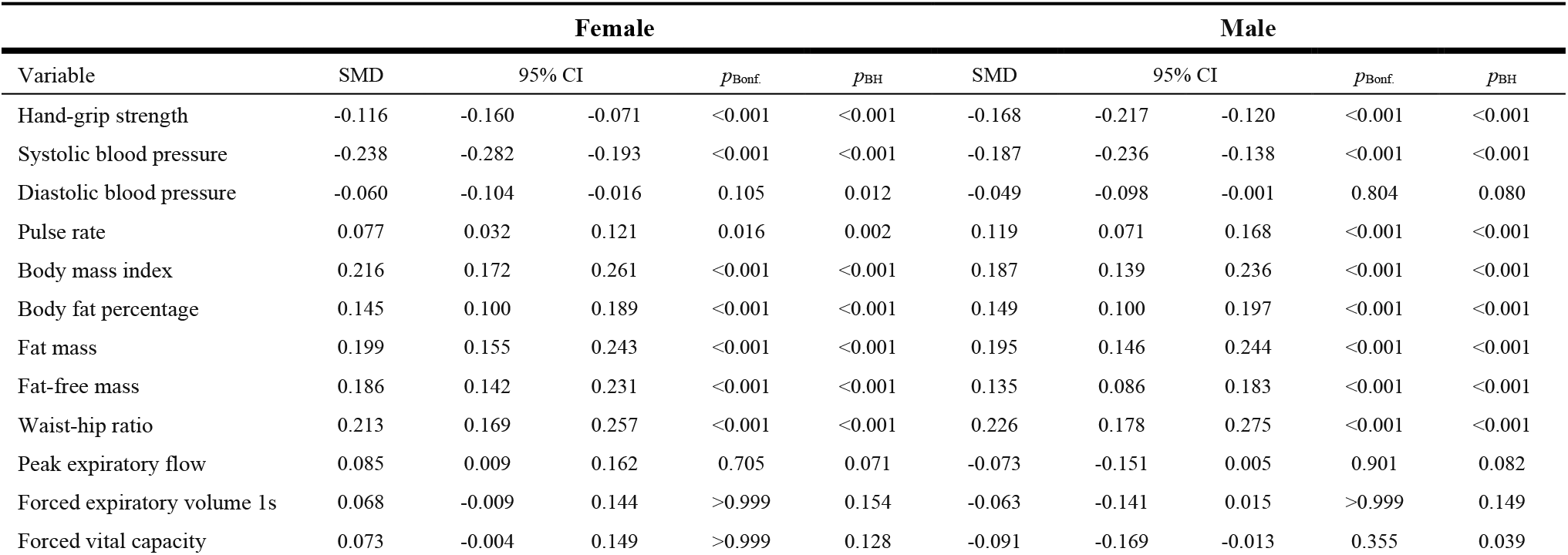
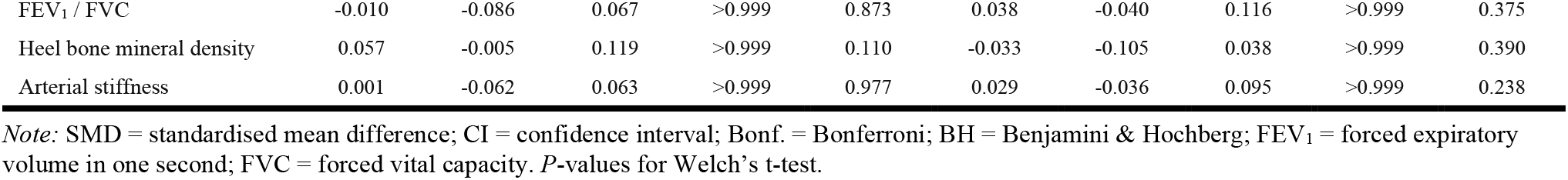
Differences in physiological measures between individuals with bipolar disorder and healthy controls

| Variable | Female |  |  |  |  | Male |  |  |  |  |
| --- | --- | --- | --- | --- | --- | --- | --- | --- | --- | --- |
| | SMD | 95% CI | | $p_{\text{Bonf.}}$ | $p_{\text{BH}}$ | SMD | 95% CI | | $p_{\text{Bonf.}}$ | $p_{\text{BH}}$ |
| Hand-grip strength | -0.116 | -0.160 | -0.071 | <0.001 | <0.001 | -0.168 | -0.217 | -0.120 | <0.001 | <0.001 |
| Systolic blood pressure | -0.238 | -0.282 | -0.193 | <0.001 | <0.001 | -0.187 | -0.236 | -0.138 | <0.001 | <0.001 |
| Diastolic blood pressure | -0.060 | -0.104 | -0.016 | 0.105 | 0.012 | -0.049 | -0.098 | -0.001 | 0.804 | 0.080 |
| Pulse rate | 0.077 | 0.032 | 0.121 | 0.016 | 0.002 | 0.119 | 0.071 | 0.168 | <0.001 | <0.001 |
| Body mass index | 0.216 | 0.172 | 0.261 | <0.001 | <0.001 | 0.187 | 0.139 | 0.236 | <0.001 | <0.001 |
| Body fat percentage | 0.145 | 0.100 | 0.189 | <0.001 | <0.001 | 0.149 | 0.100 | 0.197 | <0.001 | <0.001 |
| Fat mass | 0.199 | 0.155 | 0.243 | <0.001 | <0.001 | 0.195 | 0.146 | 0.244 | <0.001 | <0.001 |
| Fat-free mass | 0.186 | 0.142 | 0.231 | <0.001 | <0.001 | 0.135 | 0.086 | 0.183 | <0.001 | <0.001 |
| Waist-hip ratio | 0.213 | 0.169 | 0.257 | <0.001 | <0.001 | 0.226 | 0.178 | 0.275 | <0.001 | <0.001 |
| Peak expiratory flow | 0.085 | 0.009 | 0.162 | 0.705 | 0.071 | -0.073 | -0.151 | 0.005 | 0.901 | 0.082 |
| Forced expiratory volume 1s | 0.068 | -0.009 | 0.144 | >0.999 | 0.154 | -0.063 | -0.141 | 0.015 | >0.999 | 0.149 |
| Forced vital capacity | 0.073 | -0.004 | 0.149 | >0.999 | 0.128 | -0.091 | -0.169 | -0.013 | 0.355 | 0.039 |
| FEV<sub>1</sub> / FVC | -0.010 | -0.086 | 0.067 | >0.999 | 0.873 | 0.038 | -0.040 | 0.116 | >0.999 | 0.375 |
| Heel bone mineral density | 0.057 | -0.005 | 0.119 | >0.999 | 0.110 | -0.033 | -0.105 | 0.038 | >0.999 | 0.390 |
| Arterial stiffness | 0.001 | -0.062 | 0.063 | >0.999 | 0.977 | 0.029 | -0.036 | 0.095 | >0.999 | 0.238 |

Note: SMD = standardised mean difference; CI = confidence interval; Bonf. = Bonferroni; BH = Benjamini & Hochberg; FEV<sub>1</sub> = forced expiratory volume in one second; FVC = forced vital capacity. *P*-values for Welch's *t*-test.

Results from the unadjusted GAMs suggested that hand-grip strength, fat-free mass, lung function and heel bone mineral density declined with age, while systolic blood pressure, body fat percentage, fat mass (except in female cases), waist-hip ratio and arterial stiffness increased with age. Several of these results suggested non-linear relationships with age. For example, diastolic blood pressure increased until about age 55 and plateaued or decreased thereafter (Figure 3-4).

**Figure 3.**
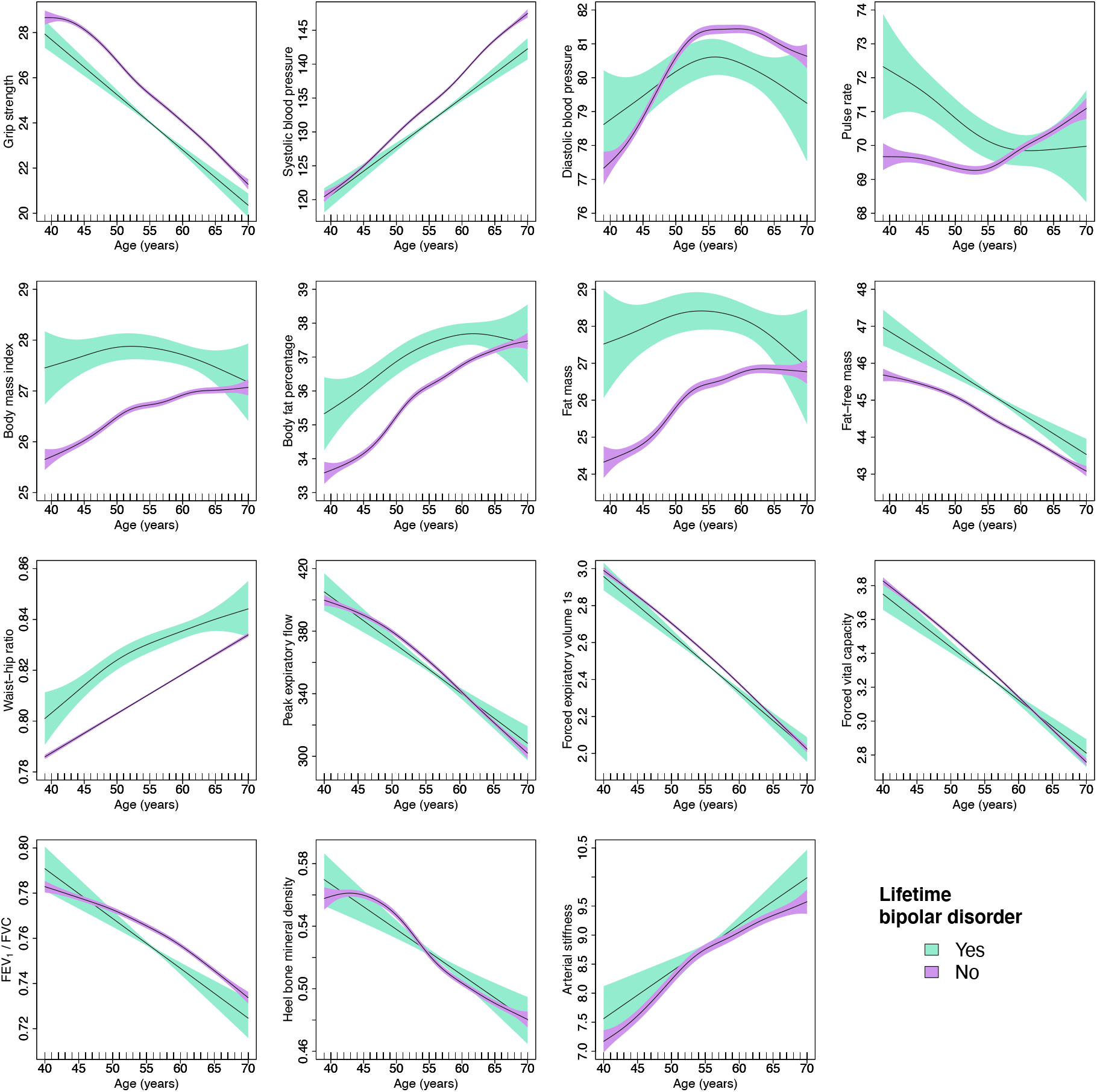
Generalised additive models of age-related changes in physiological measures in females with bipolar disorder and healthy controls. The solid lines represent physiological measures against smoothing functions of age. The shaded areas correspond to approximate 95% confidence intervals (2 standard error). FEV 1 = forced expiratory volume in one second; FVC = forced vital capacity.

**Figure 4.**
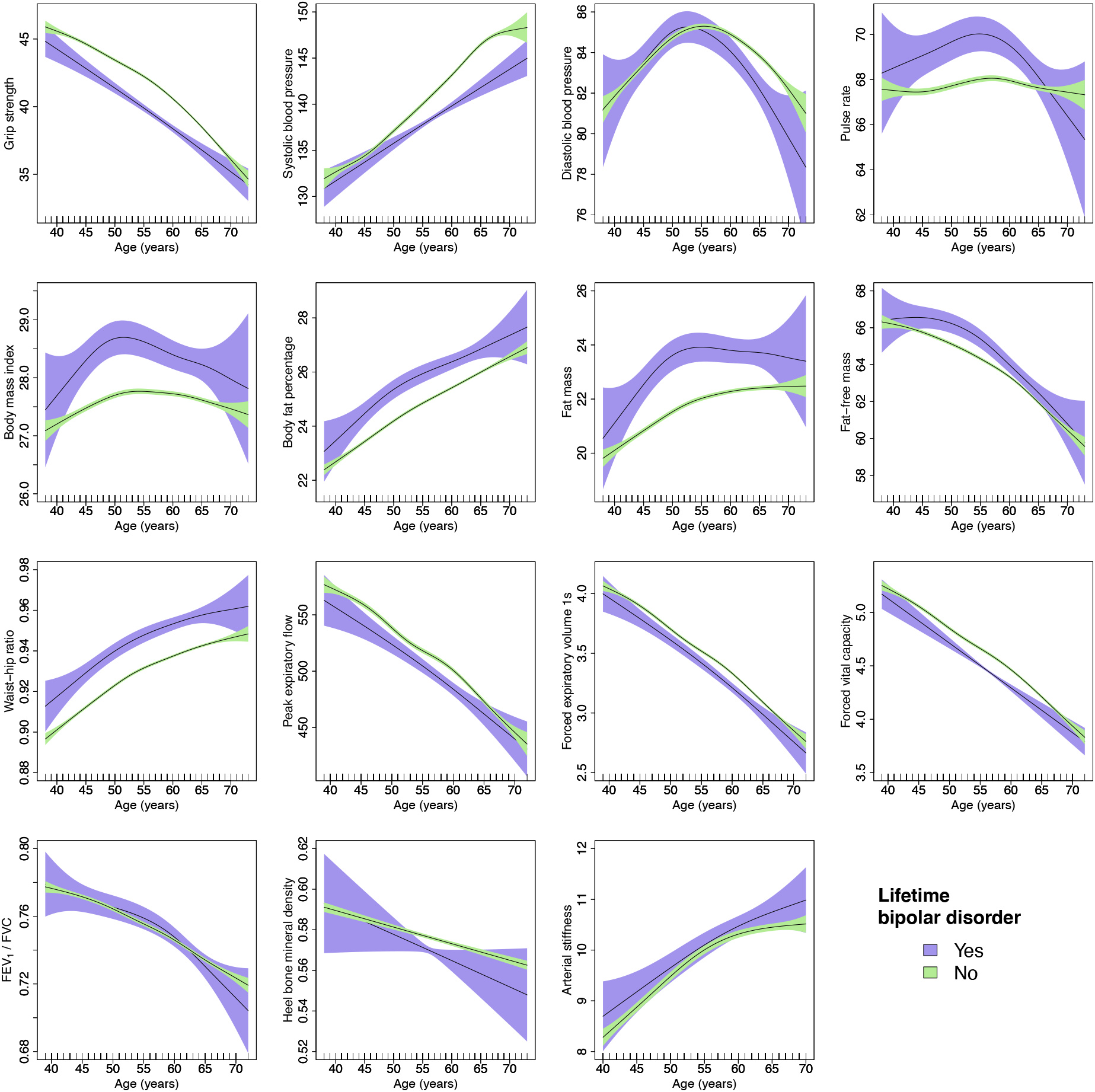
Generalised additive models of age-related changes in physiological measures in males with bipolar disorder and healthy controls. The solid lines represent physiological measures against smoothing functions of age. The shaded areas correspond to approximate 95% confidence intervals (#2 x standard error). FEV 1 = forced expiratory volume in one second; FV C = forced vital capacity.

Age-related changes in blood pressure, pulse rate, several body composition measures and, to a lesser extent, FEV1/FVC ratio differed between female cases and controls. In males, we found some evidence of differences in age-related changes in hand-grip strength, blood pressure and pulse rate, although most formal tests were not statistically significant. The other physiological measures followed comparable trajectories with age in cases and controls (Figure 5-6).

**Figure 5.**
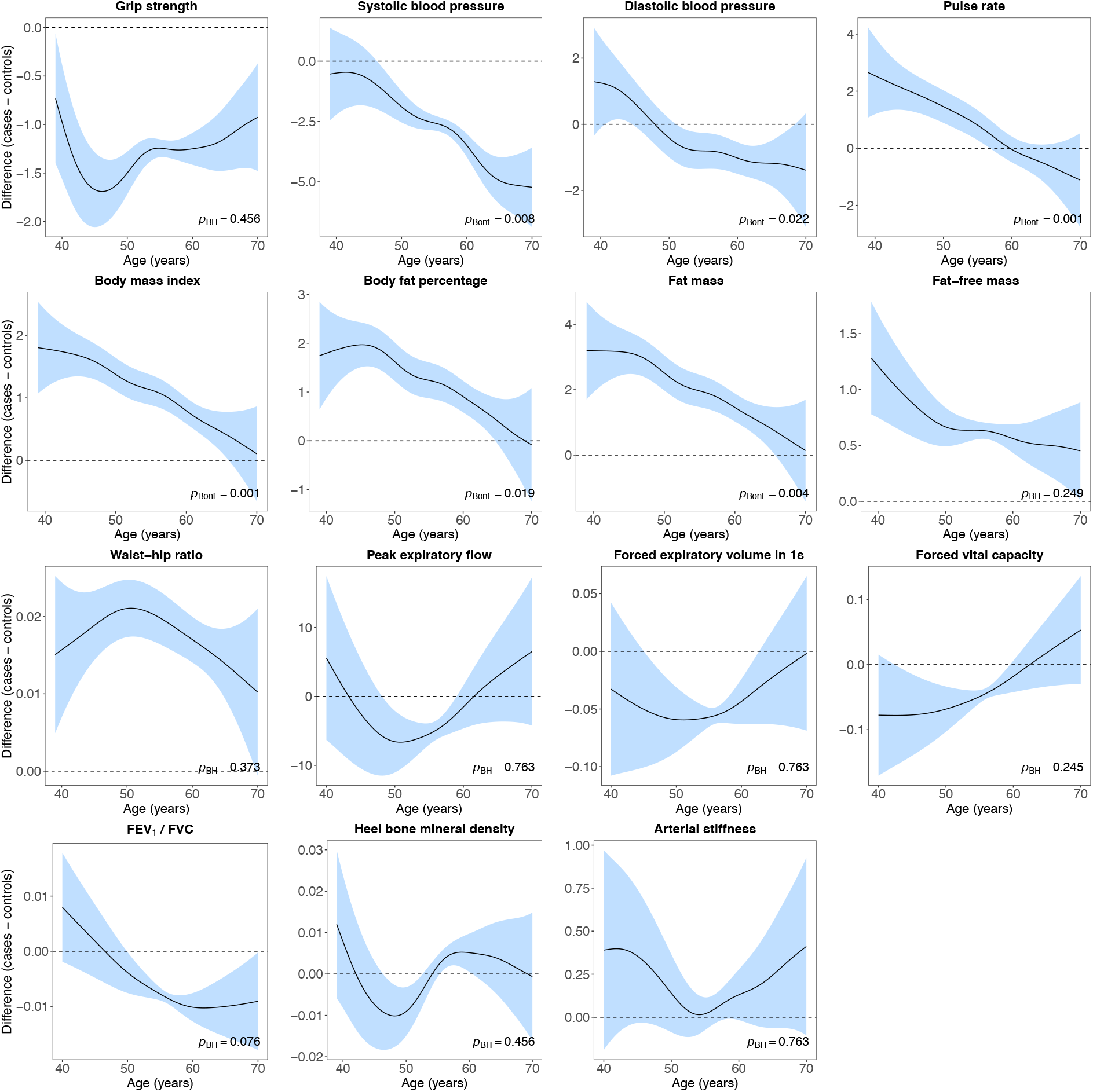
Difference smooths comparing age-related changes in physiological measures of females with bipolar disorder to healthy controls. The shaded areas correspond to approximate 95% confidence intervals (± 2 × standard error). Negative values on the y-axes correspond to lower values in females with bipolar disorder compared to healthy controls. The horizontal lines represent no difference between female cases and controls. FEV_1_ = forced expiratory volume in one second; FVC = forced vital capacity; Bonf. = Bonferroni; BH = Benjamini & Hochberg.

**Figure 6.**
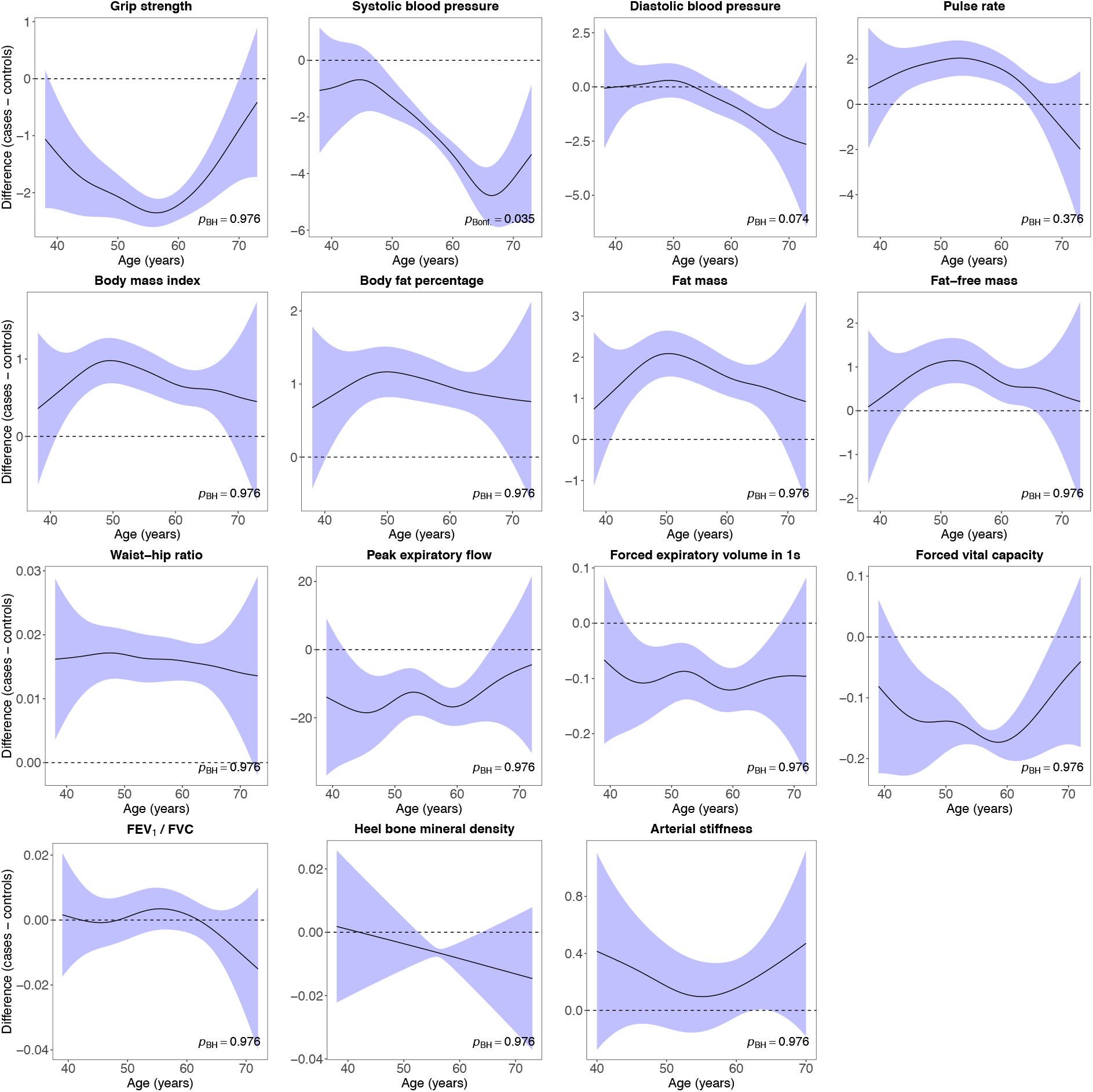
Difference smooths comparing age-related changes in physiological measures of males with bipolar disorder to healthy controls. The shaded areas correspond to approximate 95% confidence intervals (± 2 × standard error). Negative values on the y-axes correspond to lower values in males with bipolar disorder compared to healthy controls. The horizontal lines represent no difference between male cases and controls. FEV_1_ = forced expiratory volume in one second; FVC = forced vital capacity; Bonf. = Bonferroni; BH = Benjamini & Hochberg.

In females, we obtained similar results after adjustment for covariates, except that there was more overlap in the trajectories of forced vital capacity and stronger evidence of differences in the FEV_1_/FVC ratio between cases and controls (Supplement 9A). We also obtained similar results for most physiological measures in males after adjustment for covariates. However, there was no evidence of differences in age-related changes in pulse rate and none of the formal tests were statistically significant (Supplement 9B).

### Sensitivity analyses

Case-control numbers for the sensitivity analyses are presented in Supplement 10A. We retained between 21.03% to 30.84% of cases and all controls in the analyses where cases included only those with bipolar disorder according to at least two different measures. In the analyses in which we restricted our sample to participants with bipolar disorder status assessed using the MHQ, we retained between 20.09% to 43.24% of cases and between 28.31% to 35.46% of controls.

We found similar results when we applied more stringent criteria for bipolar disorder. In females, most average differences in physiological measures between cases and controls were slightly larger. However, there was no longer evidence of differences in diastolic blood pressure. Forced vital capacity was lower in female cases, although this difference was not statistically significant after multiple testing correction. In males, most differences in physiological measures were slightly larger and the difference in diastolic blood pressure remained statistically significant after multiple testing correction (Supplement 10B). Most results were also similar when we restricted the sample to individuals with bipolar disorder status assessed using the MHQ. In females, statistically significant differences were slightly larger, except for waist-hip ratio. There was no longer evidence that hand-grip strength, diastolic blood pressure or pulse rate differed between cases and controls. In males, there was no longer evidence of case-control differences in hand-grip strength or forced vital capacity. Statistically significant differences were slightly larger, except for body fat percentage and waist-hip ratio (Supplement 10B).

Most results for age-related changes in physiological measures in females were comparable to the results of our main analyses. There was some evidence that the difference smooths had different shapes for age-related changes in hand-grip strength, waist-hip ratio, peak expiratory flow, FEV_1_/FVC ratio and arterial stiffness, although these generally did not provide strong evidence of different trajectories with age. We also found stronger evidence of non-linearity in age-related differences in pulse rate between female cases according to the MHQ and healthy controls (Supplement 10C). In males, results were similar to the results of our main analyses. There was variation across analyses in the difference smooths for hand-grip strength, diastolic blood pressure, pulse rate and FEV_1_/FVC ratio, although the formal tests were generally not statistically significant after multiple testing correction. A notable difference from our main analysis was that heel bone mineral density in cases defined using the MHQ was lower at younger ages and higher at more advanced age (Supplement 10D). However, the formal test was not statistically significant.

## Discussion

This study used data from the UK Biobank to examine differences in 15 physiological measures, between males and females with bipolar disorder and healthy controls. The large sample size enabled us to highlight subtle differences between bipolar disorder cases and controls, in contrast to mixed findings from previous studies that were typically limited to clinical patient samples. We demonstrate that case-control differences for several physiological measures, most notably cardiovascular function and body composition, vary by age, and provide novel insights into the heterogeneity in ageing trajectories observed in bipolar disorder.

### Principal findings

We found small to moderate differences between bipolar disorder cases and healthy controls for hand-grip strength, blood pressure, pulse rate and body composition. There was little evidence of differences in lung function, heel bone mineral density or arterial stiffness. Case-control differences in age-related changes were most evident for cardiovascular function (in both sexes) and body composition (in females). These differences did not consistently narrow or widen with age, and some measures were higher in middle-aged cases than in controls but lower in older cases. For example, the difference in systolic blood pressure between male cases and controls was −1.3 mmHg at age 50 and widened to −4.7 mmHg at age 65. Diastolic blood pressure in female cases was 1.2 mmHg higher than in controls at age 40 and −1.2 mmHg lower at the age of 65. Body fat percentage in female cases was approximately 2% higher than in controls at age 45 and this difference narrowed with age, with no evidence of case-control differences from age 63.

### Findings in context

We observed moderate to large sex differences and evidence of age-related changes for all physiological measures. These findings are discussed in the context of other research in our previous publication13, and we focus on the bipolar disorder findings here.

### Case-control differences

Hand-grip strength was lower in cases than in controls, although not in all analyses. This finding confirms previous research. For example, a UK Biobank analysis of about 110,000 participants found slightly lower hand-grip strength in cases24. Research from Turkey showed that patients with bipolar disorder had higher rates of sarcopenia – a condition defined by loss of muscle mass and strength – than the general population25. Finally, higher hand-grip strength in Swedish adolescent males was associated with a reduced risk of bipolar disorder in adulthood26.

Cardiovascular disease and hypertension are amongst the most prevalent medical comorbidities in individuals with bipolar disorder27. However, several studies, of small sample size, found no evidence of differences in blood pressure between cases and controls28-31. We observed lower systolic blood pressure in cases, confirming a previous study in elderly euthymic outpatients32. We also found lower diastolic blood pressure in female cases, although this did not survive multiple testing in the sensitivity analyses. In males, diastolic blood pressure was lower in cases, but this difference was not statistically significant after multiple testing correction. Pulse rate was higher in cases, confirming several previous studies32-35. Previous research in newly diagnosed patients with bipolar disorder found no evidence of a statistically significant difference between cases and controls, although pulse rate was higher in patients36.

BMI, body fat percentage, fat mass, fat-free mass and waist-hip ratio were higher in male and female cases than in controls, concordant with the observation that the prevalence of self-reported obesity in US adults with bipolar disorder is higher than in controls7. Previous research on BMI has yielded mixed results, including lower BMI in cases37, higher BMI in cases28 and higher BMI only in female cases38. Other studies conducted in slightly younger populations31 or only in females39 found no evidence of differences in BMI between cases and controls. Few studies have examined differences in body fat percentage, fat mass or fat-free mass in bipolar disorder. In females, there was no evidence of differences in body fat percentage39,40, fat mass39,40 or fat-free mass40 between cases and controls. However, one study that included both sexes found higher body fat percentage in cases28. Findings from previous studies of waist-hip ratio have been mixed, including higher waist-hip ratio in cases^38^ or no difference between cases and controls^28,39^, although these studies generally had small sample sizes.

Previous research has found higher prevalence and incidence of chronic obstructive pulmonary disease in individuals with bipolar disorder compared to the general population9,41. To our knowledge, few studies have examined differences in pulmonary function between bipolar disorder cases and controls. One study found no difference in FEV1 and predicted percentage of forced vital capacity, although the sample included major depressive disorder with psychotic features and bipolar disorder42. Consistent with this study, we found no evidence of differences between female cases and controls after multiple testing correction. In males, we generally observed lower forced vital capacity in cases, although not across all analyses.

Little is known about bone health in adults with bipolar disorder^43^ and, to our knowledge, no research has examined heel bone mineral density in bipolar disorder. Research in Chinese adults found lower bone mineral density at other sites in drug-naïve bipolar disorder patients compared to healthy controls^44^. A 2019 review of three cohorts, one from the US and two from Taiwan, found higher fracture risk in individuals with bipolar disorder^45^ but included no studies that directly examined bone mineral density. In the present study, we found no evidence of differences in heel bone mineral density between cases and controls.

We found no evidence of differences in arterial stiffness between cases and controls. Few studies have examined arterial stiffness in bipolar disorder^46^ and either found no evidence of differences between cases and controls^31,47^ or age-dependent differences that suggested higher arterial stiffness in older patients with bipolar disorder compared to age-based population norms^48^.

Inconsistencies between studies could result from differences in sample characteristics such as age group or the prevalence of specific symptoms. For example, manic episodes occur less frequent in older patients while depressive episodes become more prevalent^49^. The assessment and definition of bipolar disorder, including which subtypes of bipolar disorder were examined (e.g., bipolar disorder type I, type II or both) might also contribute to differences in results.

### Case-control differences by age

To our knowledge, no previous studies have examined differences in age-related changes in hand-grip strength, blood pressure, pulse rate, body composition, lung function or heel bone mineral density between bipolar disorder cases and healthy controls, and our study provides novel insights into physiological differences in bipolar disorder across age.

We found similar rates of decline in hand-grip strength with age in bipolar disorder cases and controls. The difference in systolic blood pressure widened with age, with increasingly lower blood pressure in cases.

Although the shape of the difference smooth was similar across analyses, several of the formal tests were not statistically significant. We found that diastolic blood pressure was slightly higher in female cases until they were in their late 40s and lower thereafter, with the difference widening with age. In males, diastolic blood pressure was lower in cases from the age of 55 and this difference also widened with age. However, the formal test was not statistically significant and there was little evidence of different trajectories in the sensitivity analyses. In females, we found that differences in the pulse rate trajectories of cases and controls narrowed with age, with no evidence of higher pulse in cases from their late 50s/early 60s. This pattern was observed across most analyses. Age-related changes in pulse rate were similar in male cases and controls, although there was some evidence in the unadjusted model that differences between cases and controls were largest for those in their late 40s to late 50s.

Differences in BMI, body fat percentage and fat mass between female cases and controls narrowed with age, with no evidence of differences at age 70. A similar pattern was observed for fat-free mass, although the formal test was not statistically significant. Although the same pattern was observed across all analyses, several formal tests were not statistically significant in the sensitivity analyses. We found little evidence of differences in the trajectories of waist-hip ratio between female cases and controls, although there was some evidence of slightly larger difference in participants aged mid 40 to mid 50. In males, we found no evidence that age-related changes in body composition differed between cases and controls.

Although previous research found a higher prevalence of chronic obstructive pulmonary disease with increased age in bipolar disorder^41^, we found limited evidence that age-related changes in forced vital capacity and FEV_1_/FVC ratio differed between female cases and controls. However, most formal tests were not statistically significant. In the adjusted model, the FEV_1_/FVC ratio was higher in middle-aged cases and lower in cases at older ages. In males, we found no evidence of differences in the lung function trajectories between cases and controls.

Although the heel bone mineral density trajectories of female cases and controls overlapped, the shape of the trajectories suggested that cases followed a linear decline with age while controls followed a non-linear decline with age. We did not find evidence of differences in the trajectories of male cases and controls, but there was some evidence that the shape of the difference smooth differed between cases defined using the MHQ and controls.

To our knowledge, no previous research examined age-related changes in arterial stiffness in individuals with bipolar disorder compared to healthy controls separately for males and females. Although one previous study found higher arterial stiffness in older patients^48^, we found no evidence of differences in the trajectories of male or female cases and controls.

### Mechanisms

Several mechanisms could explain differences between bipolar disorder cases and healthy controls. For example, medications used to treat bipolar disorder may affect bone metabolism^50^ and may cause weight gain^51^. However, higher levels of obesity have also been observed in medication-naïve patients^52^, and higher prevalence of obesity or cardiovascular disease mortality in bipolar disorder were known prior to the advent of tricyclics or lithium treatment^53,54^. Differences between cases and controls could also result from risk factors pertaining to unhealthy lifestyle that are more prevalent in cases, including, for example, higher rates of smoking^55^, excessive alcohol intake^56^, substance misuse^57^ and physical inactivity^58^. These factors have broad impact across physiological systems and may also predispose individuals to comorbidities. Ineffective screening, reduced attention in treating and diagnosing physical comorbidities, misattribution of physical symptoms to mental disorder may also be relevant^59^. Finally, accelerated ageing might be a pathophysiological mechanism in bipolar disorder^60^, supported, for example, by the observation that individuals with bipolar disorder lose life-years early during the course of illness^1^.

### Limitations

As previously reported^13^, our research inevitably has limitations. Our analyses provide limited insights into the mechanism underlying our results. The cross-sectional physiological measures present uncertainty about whether these findings reflect changes in physiology due to ageing or cohort-specific differences. Previous research suggested that hand-grip strength is lower in more recent cohorts^61^, potentially resulting in underestimation of age-related decline in our study. Research on frailty indices found stability across birth cohorts^62^ as well as higher levels of frailty in younger cohorts^63,64^. The finding that some physiological differences narrowed with age could result from selection bias leading to older participants with depression in our study being healthier, relative to their age group, than younger participants, although this would not explain, for instance, why differences in blood pressure widen with age. Selection bias resulting in healthier older adults participating in the UK Biobank at higher rates could result in underestimating age-related changes. Finally, some concerns remain about the validity and reliability of the case definition for bipolar disorder. We classified cases using multiple data sources, which all have specific trade-offs. For example, mental disorders are likely underreported in linked health records, although these diagnoses may be more reliable. Self-reports might contain some level of misclassification because mental disorders are heterogenous in their presentation and have fuzzy boundaries, although participants were asked to report only diagnoses made by a doctor. Given that mental disorders fluctuate throughout the life course and we were not able to distinguish between symptomatic and euthymic cases, we examined lifetime bipolar disorder. To address potential concerns about our case definition, we conducted sensitivity analyses with more stringent criteria and found similar results. To achieve acceptable sample sizes and because not all data sources included details on bipolar disorder subtypes, we included both individuals with bipolar disorder type I and type II, which might mask potential differences between these subtypes. We might have missed some bipolar disorder type II cases in our symptom-based definition from the MHQ, as our criteria included symptom perseverance for a week, whereas the DSM-5 criteria require a minimum of four days.

### Generalisability

The response rate of the UK Biobank was low (5.5%) and compared with non-responders, participants were older, more often female, lived in less deprived neighbourhoods, engaged in healthier lifestyles and reported fewer health conditions compared with data from a nationally representative survey^65^. The UK Biobank states that “valid assessment of exposure-disease relationships are nonetheless widely generalizable and do not require participants to be representative of the population at large”^66^, in line with the observation that there is limited evidence of considerable bias due to non-participation in epidemiological research^67^. There are additional concerns about the representativeness of individuals with mental disorders because participation in research can be influenced by selection bias due to mental health^68^. Our findings do not generalise to younger and older populations and conclusions only apply to adults between the ages of 37 to 73. Additional studies that include younger participants as well as the elderly are needed.

### Implications

The primary aim of this study was to examine differences in age-related physiological changes between individuals with bipolar disorder and healthy controls. Understanding heterogeneity in ageing trajectories is of public health importance, given that variation in these measures is linked to morbidity and mortality. Nevertheless, observed associations do not imply causality. Future studies aimed at disentangling aetiology are needed to inform preventative and treatment strategies. The differences observed between cases and controls highlight the need to improve physiological health in individuals with bipolar disorder.

## Data Availability

The data used are available to all bona fide researchers for health-related research that is in the public interest, subject to an application process and approval criteria. Study materials are publicly available online at http://www.ukbiobank.ac.uk.

## Authorship contributions

CML acquired the studentship funding, interpreted the findings and critically reviewed the manuscript. JM conceived the idea of the study, acquired the data, carried out the statistical analysis, interpreted the findings, wrote the manuscript and revised the manuscript for final submission. Both authors read and approved the final manuscript. JM had full access to all data used in this study and takes responsibility for the integrity of the data and the accuracy of the data analysis.

## Acknowledgments

This research has been conducted using data from UK Biobank, a major biomedical database. This project made use of time on Rosalind HPC, funded by Guy’s & St Thomas’ Hospital NHS Trust Biomedical Research Centre (GSTT-BRC), South London & Maudsley NHS Trust Biomedical Research Centre (SLAM-BRC), and Faculty of Natural Mathematics & Science (NMS) at King’s College London.

## Ethics

Ethical approval for the UK Biobank study has been granted by the National Information Governance Board for Health and Social Care and the NHS North West Multicentre Research Ethics Committee (11/NW/0382). No project-specific ethical approval is needed. Data access permission has been granted under UK Biobank application 45514.

