## Supplement for "Age-related changes in physiology in individuals with bipolar disorder"

### Supplementary material

The following material accompanies the article "Age-related changes in physiology in individuals with bipolar disorder" by Julian Mutz and Cathryn M. Lewis

### Supplement 1. MHQ bipolar disorder criteria

| **Supplement table 1.** Case definition lifetime depression and bipolar disorder | |
|---|---|
| Variable | Definition and UK Biobank data fields |
| **Lifetime depression** (case):<br>At least one core symptom of major depressive disorder, most or all of the day on most or all days for a two-week period, with at least five non-core depressive symptoms that represent a change from usual occurring over the same timescale, with some or a lot of impairment.<br><br>No record of psychosis. | (“Ever had prolonged feelings of sadness or depression” (20446) = Yes OR “Ever had prolonged loss of interest in normal activities” (20441) = Yes)<br>AND<br>“Fraction of day affected during worst episode of depression” (20436) = Most of day or All day long<br>AND<br>“Frequency of depressed days during worst episode of depression” (20439) = “Almost every day” or “Every day”<br>AND “Impact on normal roles during worst period of depression” (20440) = “Somewhat” or “A lot”<br>AND<br>Total number of symptoms endorsed (core and others) ≥5: “Ever had prolonged feelings of sadness or depression” (core) (20446), “Ever had prolonged loss of interest in normal activities” (core) (20441), “Feelings of tiredness during worst episode of depression” (20449), “Weight change during worst episode of depression” (20536), “Did your sleep change?” (20532), “Difficulty concentrating during worst depression (20435), “Feelings of worthlessness during worst period of depression” (20450), “Thoughts of death during worst depression” (20437)<br>AND<br>**No** self-reported psychosis for “Mental health problems ever diagnosed by a professional” (20544)<br>AND<br>**No** self-reported schizophrenia for “Non-cancer illness” (20002)<br>AND<br>**No** ICD-10 code for “schizophrenia, schizotypal and delusional disorders” (F20-F29)<br>AND<br>**No** primary care record of psychosis |
| **Lifetime bipolar disorder** (case):<br>Meeting criteria for lifetime depression reported above, having had a period of mania/excitability or extreme irritability for a week or more, and endorsing at least three other features (four if no period of mania/excitability).<br><br>No record of psychosis. | Lifetime depression (case) as reported above<br>AND<br>(“Ever had period of mania/excitability” (20501) = Yes OR “Ever had period extreme irritability” (20502) = Yes)<br>AND<br>Three (four if “Ever had period of mania/excitability” (20501) = No) features endorsed from “Manifestations of mania or irritability” (20548): (1) I was more talkative than usual; (2) I was more restless than usual; (3) My thoughts were racing; (4) I needed less sleep than usual; (5) I was more creative or had more ideas than usual; (6) I was easily distracted; (7) I was more confident than usual; (8) I was more active than usual<br>AND<br>“Longest period of mania or irritability” (20492) = A week or more.<br>AND<br>**No** self-reported psychosis for “Mental health problems ever diagnosed by a professional” (20544)<br>AND<br>**No** self-reported schizophrenia for “Non-cancer illness” (20002)<br>AND<br>**No** ICD-10 code for “schizophrenia, schizotypal and delusional disorders” (F20-F29)<br>AND<br>**No** primary care record of psychosis |
| *Note:* Criteria for lifetime depression and bipolar disorder adapted from Davis et al. (2020), doi: 10.1192/bjo.2019.100. ICD-10 = International Classification of Diseases, Tenth Revision; MHQ = mental health questionnaire. | |

#### Supplement 2. ICD-10 codes for bipolar disorder

**Manic episode**

- F30.0 Hypomania
- F30.1 Mania without psychotic symptoms
- F30.2 Mania with psychotic symptoms
- F30.8 Other manic episodes
- F30.9 Manic episode, unspecified

**Bipolar affective disorder**

- F31.0 Bipolar affective disorder, current episode hypomanic
- F31.1 Bipolar affective disorder, current episode manic without psychotic symptoms
- F31.2 Bipolar affective disorder, current episode manic with psychotic symptoms
- F31.3 Bipolar affective disorder, current episode mild or moderate depression
- F31.4 Bipolar affective disorder, current episode severe depression without psychotic symptoms
- F31.5 Bipolar affective disorder, current episode severe depression with psychotic symptoms
- F31.6 Bipolar affective disorder, current episode mixed
- F31.7 Bipolar affective disorder, currently in remission
- F31.8 Other bipolar affective disorders
- F31.9 Bipolar affective disorder, unspecified

Supplement 3. Smith et al. bipolar disorder criteria

**Bipolar disorder adapted from** Smith et al. (2013).

**1. Probable bipolar disorder (type I):**
4642 ever manic/hyper 2 days OR 4653 ever irritable/argumentative for 2 days;
plus at least 3 from 6156.01 (more active), 6156.02 (more talkative), 6156.03 (needed less sleep), and 6156.04 (more creative/more ideas);
plus 5663 duration of a week or more;
plus 5674 needed treatment or caused problems at work.

**2. Probable bipolar disorder (type II):**
4642 ever manic/hyper 2 days OR 4653 ever irritable/argumentative for 2 days;
plus at least 3 from 6156.01 (more active), 6156.02 (more talkative), 6156.03 (needed less sleep), and 6156.04 (more creative/more ideas);
plus 5663 duration of a week or more.

### Supplement 4. Psychotropic medication codes

**Supplement table 2.** Psychotropic medication codes

| UK Biobank code | Drug name |
|---|---|
| 1140879616 | Amitriptyline |
| 1140921600 | Citalopram |
| 1140879540 | Fluoxetine |
| 1140867878 | Sertraline |
| 1140916282 | Venlafaxine |
| 1140909806 | Dosulepin |
| 1140867888 | Paroxetine |
| 1141152732 | Mirtazapine |
| 1141180212 | Escitalopram |
| 1140879634 | Trazodone |
| 1140867876 | Prozac |
| 1140882236 | Seroxat |
| 1141190158 | Cipralex |
| 1141200564 | Duloxetine |
| 1140867726 | Lofepramine |
| 1140879620 | Clomipramine |
| 1140867818 | Nortriptyline |
| 1140879630 | Imipramine |
| 1140879628 | Dothiepin |
| 1141151946 | Cipramil |
| 1140867948 | Amitriptyline |
| 1140867624 | Prothiaden |
| 1140867756 | Trimipramine |
| 1140867884 | Lustral |
| 1141151978 | Reboxetine |
| 1141152736 | Zispin |
| 1141201834 | Cymbalta |
| 1140867690 | Anafranil |
| 1140867640 | Doxepin |
| 1140867920 | Moclobemide |
| 1140867850 | Phenelzine |
| 1140879544 | Fluvoxamine |
| 1141200570 | Yentreve |
| 1140867934 | Triptafen |
| 1140867758 | Surmontil |
| 1140867914 | Tranylcypromine |
| 1140867820 | Allegron |
| 1141151982 | Edronax |
| 1140882244 | Molipaxin |
| 1140879556 | Mianserin |
| 1140867852 | Nardil |
| 1140867860 | Faverin |
| 1140917460 | Nefazodone |
| 1140867938 | Amitriptyline+Chlordiazepoxide |
| 1140867856 | Isocarboxazid |
| 1140867922 | Manerix |
| 1140910820 | Maoi |
| 1140882312 | Sinequan |
| 1140867944 | Tranylcypromine+Trifluoperazine |
| 1140867784 | Ludiomil |
| 1140867812 | Norval |
| 1140867668 | Tryptizol |
| 1140867940* | Fluphenazine hydrochloride+Nortriptyline 1.5mg/30mg tablet |
| 1140867942* | Fluphenazine hcl+Nortriptyline 500micrograms/10mg tablet |
| 1140928916 | Olanzapine |
| 1141152848 | Quetiapine |
| 1140867444 | Risperidone |
| 1140879658 | Chlorpromazine |
| 1140868120 | Trifluoperazine |
| 1141153490 | Amisulpride |
| 1140867304 | Sulpiride |
| 1141152860 | Seroquel |
| 1140867168 | Haloperidol |
| 1141195974 | Aripiprazole |
| 1140867244 | Stelazine |
| 1140867152 | Depixol |
| 1140909800 | Flupentixol |
| 1140867420 | Clozapine |
| 1140879746 | Promazine |
| 1141177762 | Risperdal |

| | |
|---|---|
| 1140867456 | Modecate |
| 1140867952 | Fluanxol |
| 1140867150 | Flupenthixol |
| 1141167976 | Zyprexa |
| 1140882100 | Zuclopenthixol |
| 1140867342 | Clopixol |
| 1140863416 | Largactil |
| 1141202024 | Abilify |
| 1140882098 | Fluphenazine |
| 1140867184 | Haldol |
| 1140867092 | Serenace |
| 1140882320 | Clozaril |
| 1140910358 | Cpz |
| 1140867208 | Perphenazine |
| 1140909802 | Levomepromazine |
| 1140867134 | Pericyazine |
| 1140867306 | Dolmatil |
| 1140867210 | Fentazin |
| 1140867398 | Fluphenazine |
| 1140867078 | Benperidol |
| 1140867218 | Pimozide |
| 1141201792 | Zaponex |
| 1141200458 | Denzapine |
| 1140867136 | Neulactil |
| 1140879750 | Thioridazine |
| 1140867180 | Dozic |
| 1140867546 | Fluspirilene |
| 1140928260 | Panadeine |
| 1140927956 | Sertindole |
| 1140867490 | Lithium product |
| 1140867494 | Camcolit 250 tablet |
| 1140867498 | Liskonum 450mg m/r tablet |
| 1140867500 | Phasal 300mg m/r tablet |
| 1140867504 | Priadel 200mg m/r tablet |
| 1140867518 | Litarex 564mg m/r tablet |
| 1140867520 | Li-liquid 5.4mmol/5ml oral solution |

*Note:* Adapted from Davis et al. (2019), doi: 10.1002/mpr.1796. *medication code not included in Davis et al. (2019).

### Supplement 5. Physiological measures

Description of physiological measures reproduced from Mutz et al. (2021)[1].

**Physiological measures**
Data on physiological measures were collected by certified healthcare technicians or nurses using a direct data entry system. Participants were asked to remove any outer garments, shoes, socks or tights. The assessment lasted 10-15 minutes.

**Hand-grip strength**
Hand-grip strength in whole kilogram force units was measured using a Jamar J00105 hydraulic hand dynamometer (measurement range 0-90 kg). Participants were asked to sit upright in a chair and place their forearms on armrests. The dynamometer handle was set to the second incremental slot or, in participants with very large hands, the third slot. Participants kept their elbow adjacent to their side and bent to a 90° angle with their thumb facing upward. A maximal score was obtained from each participant's right and left hand. We used the maximal grip strength of the participant's self-reported dominant hand. If no data on handedness were available, we used the highest value of both hands. This variable has been used previously in UK Biobank research[2], although other studies have used the highest value of left and right hand[3] or calculated the average grip strength from both hands[4]. We used absolute units because these are the simplest to use in risk assessment. A previous UK Biobank study found no evidence of differences in mortality or disease incidence prediction when grip strength was expressed in absolute terms (kilograms) compared to when expressed relative to anthropometric traits (height, weight, fat-free mass or BMI)[4].

**Body composition**
Weight measurements were obtained with a Tanita BC-418 MA body composition analyser, or, in individuals with pacemaker and females who reported that they were pregnant, using a manual scale. Standing height was measured using a Seca 202 height measure. BMI was calculated as weight divided by standing height squared ($kg/m^2$). Waist and hip circumference in cm were measured using a Wessex non-stretchable sprung tape. Waist-to-hip ratio was calculated by dividing waist circumference by hip circumference. Whole body fat mass and fat-free mass in kg and body fat percentage (measurement range 1-75%) were estimated by electrical bio-impedance using a Tanita BC-418 MA.

**Pulmonary function**
Volumetric measures of lung function were quantified using breath spirometry with a Vitalograph Pneumotrac 6800. Participants were asked to record two to three blows, each lasting for at least six seconds, within a period of approximately six minutes. The reproducibility of the first two blows was automatically compared and, if acceptable (defined as a <5% difference in forced vital capacity (FVC) and forced expiratory volume in one second ($FEV_1$)), a third blow was not required. FVC in litres describes the maximum amount of air that can be exhaled when blowing out as fast as possible after a deep breath. $FEV_1$ in litres describes the amount of air that can be exhaled in one second when blowing out as fast as possible. We used the derived best measure for both variables which was the maximum value from reproducible spirograms, in line with previous research[5]. Given that these measures are affected by several factors unrelated to pulmonary function (e.g., effort and body size), we also calculated the ratio of $FEV_1$ to FVC. Peak expiratory flow (PEF) in litres per minute represents a person's maximum speed of expiration. PEF is determined by physiological factors such as lung volume and elasticity or expiratory muscle strength. It is used for monitoring asthma and diagnosing chronic obstructive pulmonary disease. We calculated the average of all available readings. Spirometry was not performed in participants who had any of the following contra-indications: chest infection in the last month (i.e., influenza, bronchitis, severe cold, pneumonia); history of a detached retina; heart attack or surgery to eyes, chest or abdomen in the last three months; history of a collapsed lung; pregnancy in the 1st or 3rd trimester; currently on medication for tuberculosis.

**Heel bone mineral density**
Heel bone mineral density was estimated by quantitative ultrasound assessment of the calcaneus using a Sahara Clinical Bone Sonometer. Participants were asked to sit with their back straight and had their left heel measured first. Measurement of both heels was performed only during later stages of the baseline assessment. Measures of speed of sound in metres per second and broadband ultrasound attenuation (decibels/megahertz) were combined into a quantitative ultrasound index. From this an estimate of heel bone mineral density in grams per $cm^2$ was derived based on the assumption that sound waves travel differently through denser bones.

**Cardiovascular measures**

Blood pressure, pulse rate and arterial stiffness (pulse wave velocity) were measured by trained nurses during a separate stage of the baseline assessment.

**Blood pressure**

Seated systolic and diastolic resting blood pressure in millimetres of mercury (mmHg) was measured twice using an Omron 705 IT digital blood pressure monitor (measurement range 0-255 mmHg) following standard procedures. Participants were asked to loosen or remove any restrictive clothing and put their arm on a desktop. Measurements were taken from the left upper arm or, if impractical, from the right arm. If there was a problem with the measurement, it was repeated. Three different cuff sizes were available and a Seca tape was used to determine the circumference of the midpoint of the upper arm. If the largest cuff size was too small or the digital blood pressure monitor did not produce a reading, a manual sphygmomanometer was used in conjunction with a stethoscope. A second measurement was taken at least one minute after the first measurement. We used an average of the two readings to reduce potential measurement error.

**Pulse rate**

Resting pulse rate in beats per minute was recorded during the blood pressure measurements using the Omron 705 IT device or, exceptionally, a manual sphygmomanometer. We used an average of the two readings to reduce potential measurement error.

**Arterial stiffness**

Resting pulse wave velocity was measured non-invasively using finger photoplethysmography with a PulseTrace PCA2 infra-red sensor. The pulse waveform was recorded over a period of 10-15 seconds, with the sensor clipped to the end of the index finger of the non-dominant hand while the participant was sitting. If the waveform did not fill at least 2/3 of the display of the device, or did not stabilize within one minute, the measurement was repeated on a larger finger or on the thumb. The shape of the waveform is directly related to the time it takes for the pulse wave to travel through the arterial tree in the lower body and to be reflected back to the finger. The time between the peaks of the waveform (the pulse wave peak-to-peak time, i.e., the difference between the peak values of direct and reflected components) was divided by the participant's height to obtain the arterial stiffness index in metres per second. A higher score on the index represents stiffer arteries. The method has been externally validated and is highly correlated with the gold-standard carotid-femoral pulse wave velocity[6].

### Supplement 6. Antihypertensive medication codes

**Supplement table 3.** Antihypertensive medication codes

| UK Biobank code | Drug name |
|---|---|
| 1140861848 | Colestid 5g/sachet granules |
| 1140861922 | Lipid lowering drug |
| 1140861924 | Bezafibrate |
| 1140861926 | Bezalip 200mg tablet |
| 1140861936 | Questran 4g/sachet powder |
| 1140861944 | Clofibrate |
| 1140861954 | Fenofibrate |
| 1140861958 | Simvastatin |
| 1140861970 | Lipostat 10mg tablet |
| 1140862026 | Ciprofibrate |
| 1140864592 | Lescol 20mg capsule |
| 1140881748 | Zocor 10mg tablet |
| 1140888590 | Colestipol |
| 1140888594 | Fluvastatin |
| 1140888648 | Pravastatin |
| 1140909780 | Colestyramine |
| 1141146138 | Lipitor 10mg tablet |
| 1141146234 | Atorvastatin |
| 1141157260 | Bezafibrate product |
| 1141162544 | Lipantil micro 67mg capsule |
| 1141172214 | Supralip 160mg m/r tablet |
| 1141180734 | Colestyramine product |
| 1141188146 | Simvador 10mg tablet |
| 1141192410 | Rosuvastatin |
| 1141192414 | Crestor 10mg tablet |
| 1141192736 | Ezetimibe |
| 1141192740 | Ezetrol 10mg tablet |
| 1141195196 | Ranzolont 10mg tablet |
| 1141200040 | Zocor heart-pro 10mg tablet |

### Supplement 7. Case numbers by data sources

**Supplement table 4.** Bipolar disorder case numbers from various data sources

| Dataset | Data source | | | | | |
|---|---|---|---|---|---|---|
| | MHQ symptom-based definition | MHQ single-item question | Baseline nurse-led interview | Hospital inpatient record | Smith et al. criteria | Primary care record |
| Main dataset | 1181 | 506 | 778 | 770 | 1154 | 633 |
| Lung function | 438 | 205 | 296 | 262 | 386 | 236 |
| Bone mineral density | 739 | 326 | 501 | 506 | NA | 410 |
| Arterial stiffness | 446 | 179 | 297 | 283 | 1179 | 242 |

*Note:* Numbers are not mutually exclusive. MHQ = mental health questionnaire; NA = not available.

### Supplement 8. Sample characteristics

**Supplement table 5.** Sample characteristics

| | | Female | | Male | |
|---|---|---|---|---|---|
| | Overall (N=502521) | Healthy control (N=132900) | Bipolar disorder (N=1984) | Healthy control (N=135933) | Bipolar disorder (N=1645) |
| **Age** | | | | | |
| Mean (SD) | 56.53 (8.10) | 55.81 (8.02) | 53.53 (7.72) | 56.31 (8.25) | 55.04 (8.11) |
| **Ethnicity** | | | | | |
| White | 472711 (94.1%) | 126345 (95.1%) | 1872 (94.4%) | 129619 (95.4%) | 1556 (94.6%) |
| Mixed-race | 2958 (0.6%) | 871 (0.7%) | 21 (1.1%) | 613 (0.5%) | 16 (1.0%) |
| Asian | 8061 (1.6%) | 2179 (1.6%) | 33 (1.7%) | 1811 (1.3%) | 20 (1.2%) |
| Black | 9882 (2.0%) | 1883 (1.4%) | 33 (1.7%) | 2574 (1.9%) | 29 (1.8%) |
| Chinese | 1574 (0.3%) | 554 (0.4%) | 4 (0.2%) | 368 (0.3%) | 4 (0.2%) |
| Other | 4558 (0.9%) | 1068 (0.8%) | 21 (1.1%) | 948 (0.7%) | 20 (1.2%) |
| Prefer not to answer | 1662 (0.3%) | | | | |
| Do not know | 217 (0.0%) | | | | |
| Missing | 898 (0.2%) | | | | |
| **Household income**[1] | | | | | |
| Very low | 97205 (19.3%) | 35212 (26.5%) | 489 (24.6%) | 37980 (27.9%) | 409 (24.9%) |
| Low | 108177 (21.5%) | 28048 (21.1%) | 591 (29.8%) | 22647 (16.7%) | 450 (27.4%) |
| Middle | 110774 (22.0%) | 34783 (26.2%) | 516 (26.0%) | 32487 (23.9%) | 377 (22.9%) |
| High | 86269 (17.2%) | 27319 (20.6%) | 304 (15.3%) | 33260 (24.5%) | 332 (20.2%) |
| Very high | 22930 (4.6%) | 7538 (5.7%) | 84 (4.2%) | 9559 (7.0%) | 77 (4.7%) |
| Prefer not to answer | 49848 (9.9%) | | | | |
| Do not know | 21305 (4.2%) | | | | |
| Missing | 6013 (1.2%) | | | | |
| **Walking**[2] | | | | | |
| Mean (SD) | 5.39 (1.93) | 5.48 (1.85) | 5.43 (1.99) | 5.31 (1.99) | 5.33 (2.09) |
| Prefer not to answer | 979 (0.2%) | | | | |
| Unable to walk | 1929 (0.4%) | | | | |
| Do not know | 6687 (1.3%) | | | | |
| Missing | 874 (0.2%) | | | | |
| **Moderate activity**[2] | | | | | |
| Mean (SD) | 3.63 (2.33) | 3.62 (2.31) | 3.66 (2.42) | 3.59 (2.31) | 3.63 (2.44) |
| Prefer not to answer | 2273 (0.5%) | | | | |
| Do not know | 24120 (4.8%) | | | | |
| Missing | 878 (0.2%) | | | | |
| **Vigorous activity**[2] | | | | | |
| Mean (SD) | 1.84 (1.96) | 1.73 (1.85) | 1.74 (1.97) | 2.10 (2.02) | 2.04 (2.11) |
| Prefer not to answer | 4116 (0.8%) | | | | |
| Do not know | 22582 (4.5%) | | | | |
| Missing | 878 (0.2%) | | | | |
| **Smoking status** | | | | | |
| Never | 273528 (54.4%) | 82219 (61.9%) | 976 (49.2%) | 70308 (51.7%) | 664 (40.4%) |
| Former | 173064 (34.4%) | 40758 (30.7%) | 653 (32.9%) | 50780 (37.4%) | 636 (38.7%) |
| Current | 52979 (10.5%) | 9923 (7.5%) | 355 (17.9%) | 14845 (10.9%) | 345 (21.0%) |
| Prefer not to answer | 2059 (0.4%) | | | | |
| Missing | 891 (0.2%) | | | | |
| **Alcohol intake frequency** | | | | | |
| Never | 40645 (8.1%) | 35495 (26.7%) | 463 (23.3%) | 35392 (26.0%) | 366 (22.2%) |
| Special occasions | 58011 (11.5%) | 9803 (7.4%) | 232 (11.7%) | 6519 (4.8%) | 190 (11.6%) |
| 1-3/month | 55856 (11.1%) | 17528 (13.2%) | 340 (17.1%) | 8562 (6.3%) | 130 (7.9%) |
| 1-2/week | 129294 (25.7%) | 17129 (12.9%) | 292 (14.7%) | 11634 (8.6%) | 163 (9.9%) |
| 3-4/week | 115443 (23.0%) | 30146 (22.7%) | 329 (16.6%) | 37873 (27.9%) | 372 (22.6%) |
| Daily/almost daily | 101770 (20.3%) | 22799 (17.2%) | 328 (16.5%) | 35953 (26.4%) | 424 (25.8%) |
| Prefer not to answer | 605 (0.1%) | | | | |
| Missing | 897 (0.2%) | | | | |
| **Sleep duration** | | | | | |
| Mean (SD) | 7.15 (1.11) | 7.18 (1.00) | 7.29 (1.37) | 7.12 (0.99) | 7.13 (1.40) |
| Prefer not to answer | 386 (0.1%) | | | | |
| Do not know | 2943 (0.6%) | | | | |
| Missing | 887 (0.2%) | | | | |
| **Antihypertensive use** | | | | | |
| Yes | 85084 (16.93%) | 13317 (10.0%) | 216 (10.9%) | 26363 (19.4%) | 361 (21.9%) |
| No | 416575 (82.90%) | 119583 (90.0%) | 1768 (89.1%) | 109570 (80.6%) | 1284 (78.1%) |
| Missing | 862 (0.17%) | | | | |

*Note:* Descriptive statistics for covariates based on main dataset *N*=272 462. [1]Annual household income groups: very low (<£18 000), low (£18 000–30 999), middle (£31 000–51 999), high (£52 000–100 000) and very high (>£100 000). [2]number of days per week engaging in these activities for 10+ minutes continuously.

### Supplement 9. Adjusted GAMs

#### 9A. Age-related changes in females

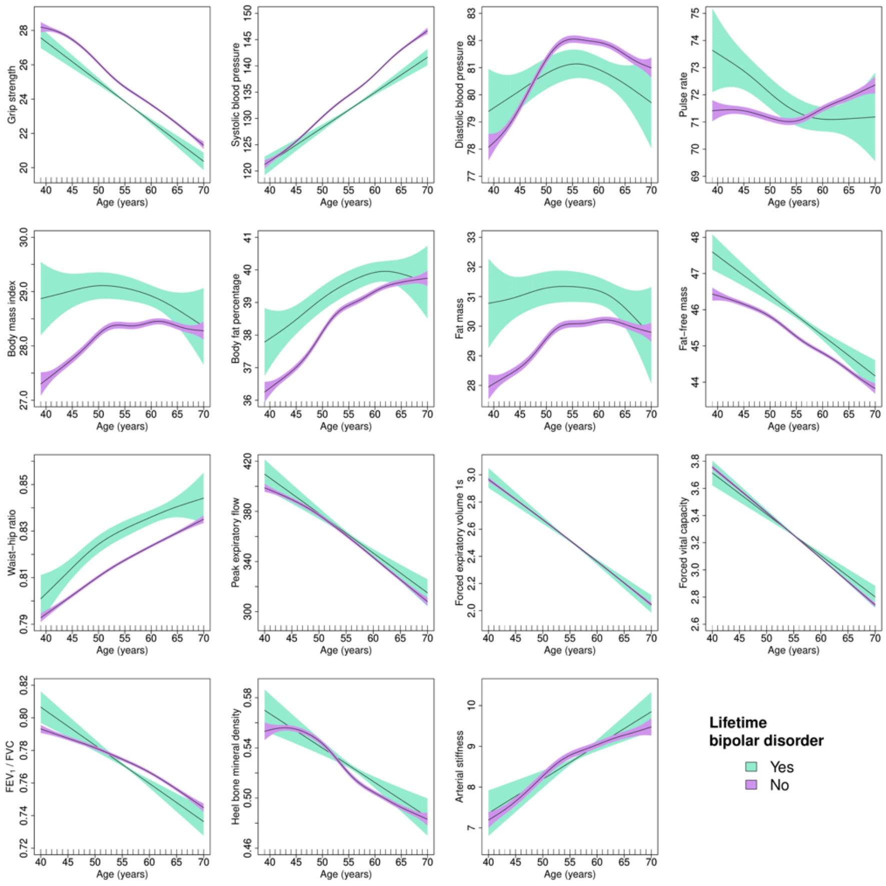

**Supplement figure 1.** Adjusted generalised additive models of age-related changes in physiological measures in females with bipolar disorder and healthy controls. Models were adjusted for ethnicity (except lung function), gross annual household income, physical activity, smoking status, alcohol intake frequency, sleep duration and, for cardiovascular measures, current use of antihypertensive medications. The solid lines represent physiological measures against smoothing functions of age. The shaded areas correspond to approximate 95% confidence intervals (± 2 × standard error). $FEV_1$ = forced expiratory volume in one second; FVC = forced vital capacity.

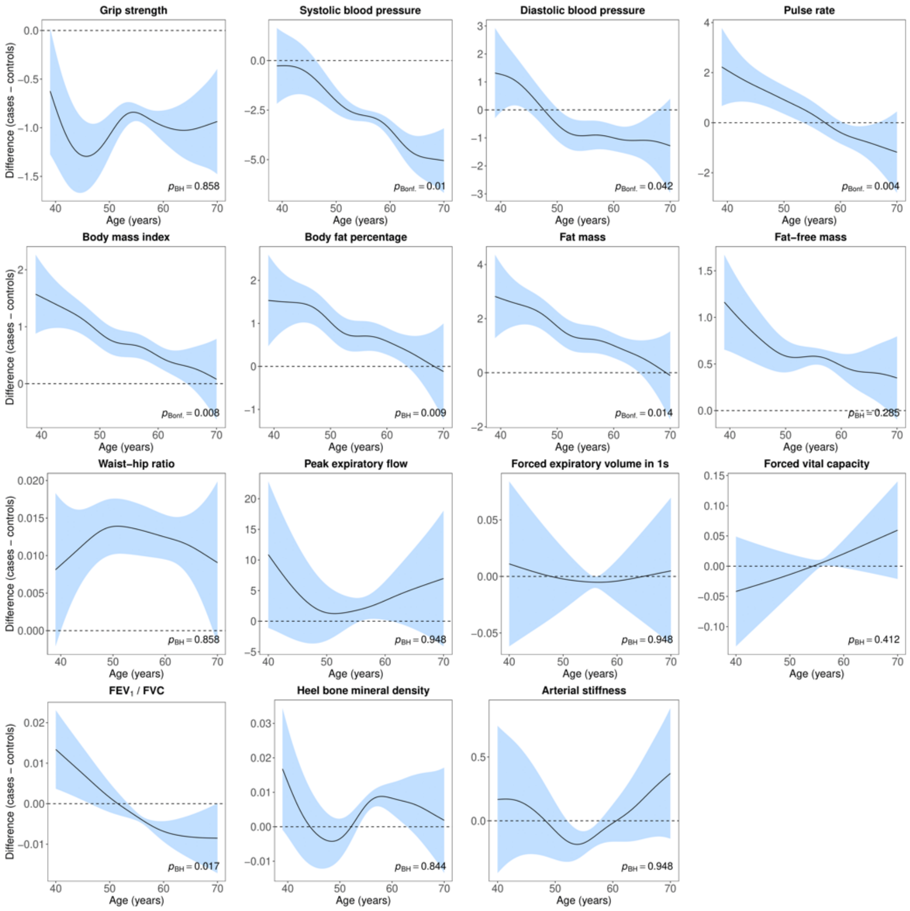

**Supplement figure 2.** Difference smooths comparing age-related changes in physiological measures of females with bipolar disorder to healthy controls. Models were adjusted for ethnicity (except lung function), gross annual household income, physical activity, smoking status, alcohol intake frequency, sleep duration and, for cardiovascular measures, current use of antihypertensive medications. The shaded areas correspond to approximate 95% confidence intervals (± 2 × standard error). Negative values on the y-axes correspond to lower values in females with bipolar disorder compared to healthy controls. The horizontal lines represent no difference between female cases and controls. $FEV_1$ = forced expiratory volume in one second; FVC = forced vital capacity; Bonf. = Bonferroni; BH = Benjamini & Hochberg.

9B. Age-related changes in males

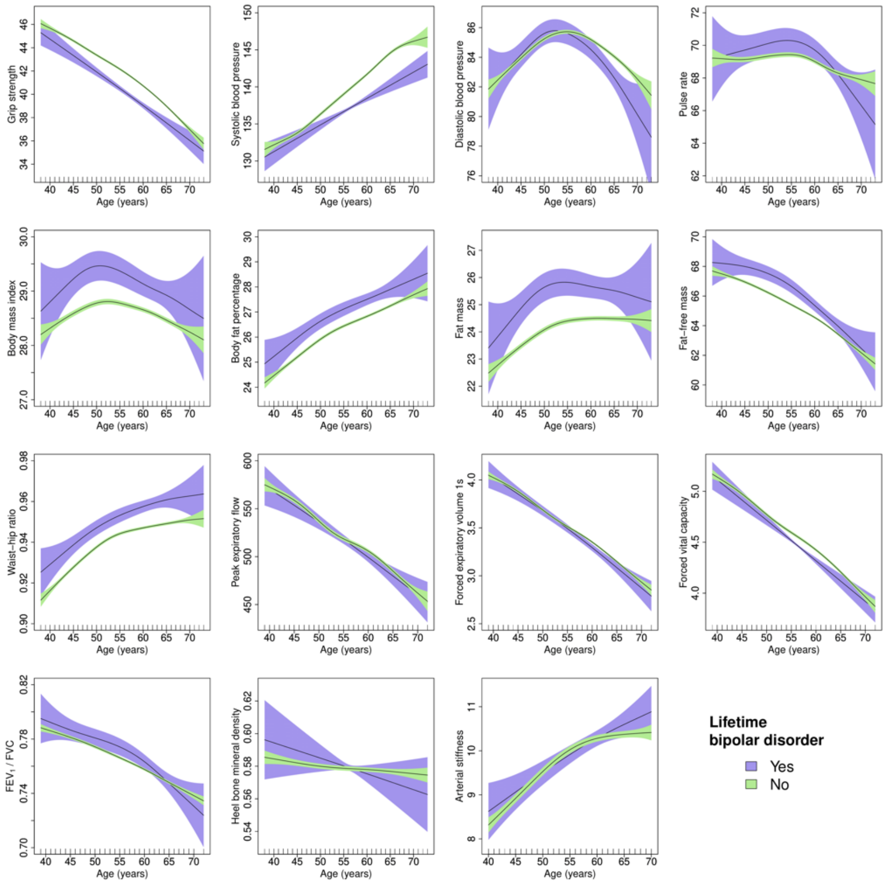

**Supplement figure 3.** Adjusted generalised additive models of age-related changes in physiological measures in males with bipolar disorder and healthy controls. Models were adjusted for ethnicity (except lung function), gross annual household income, physical activity, smoking status, alcohol intake frequency, sleep duration and, for cardiovascular measures, current use of antihypertensive medications. The solid lines represent physiological measures against smoothing functions of age. The shaded areas correspond to approximate 95% confidence intervals (± 2 × standard error). $FEV_1$ = forced expiratory volume in one second; FVC = forced vital capacity.

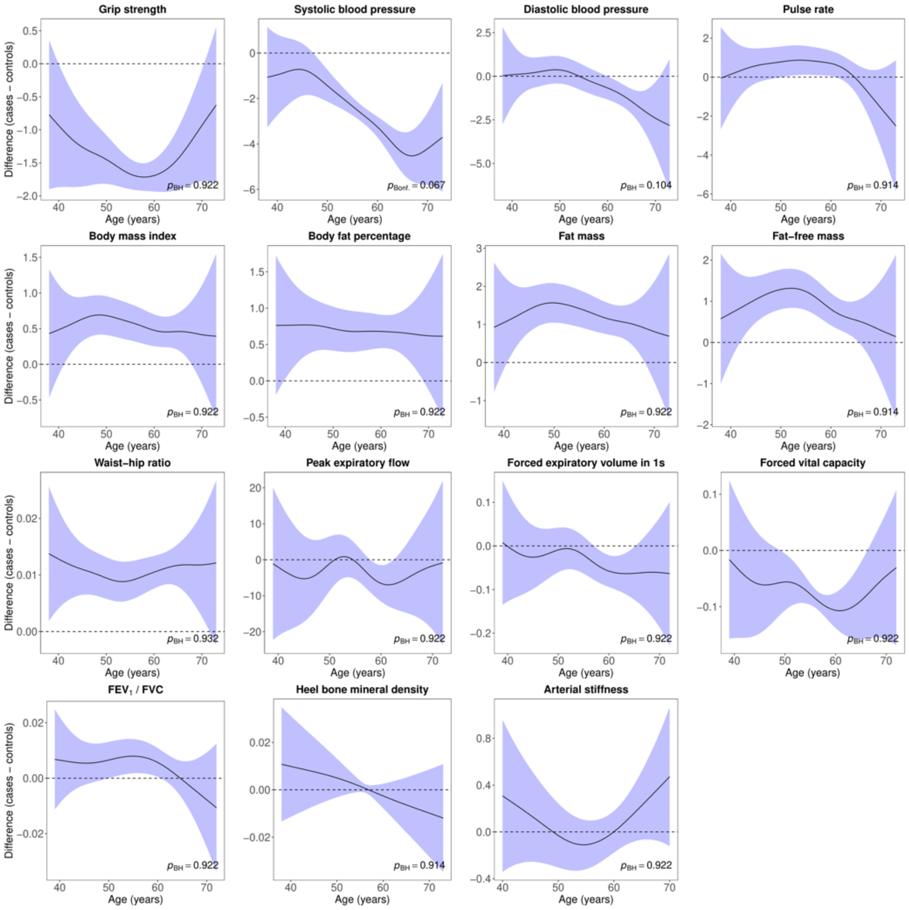

**Supplement figure 4.** Difference smooths comparing age-related changes in physiological measures of males with bipolar disorder to healthy controls. Models were adjusted for ethnicity (except lung function), gross annual household income, physical activity, smoking status, alcohol intake frequency, sleep duration and, for cardiovascular measures, current use of antihypertensive medications. The shaded areas correspond to approximate 95% confidence intervals (± 2 × standard error). Negative values on the y-axes correspond to lower values in males with bipolar disorder compared to healthy controls. The horizontal lines represent no difference between male cases and controls. $FEV_1$ = forced expiratory volume in one second; FVC = forced vital capacity; Bonf. = Bonferroni; BH = Benjamini & Hochberg.

### Supplement 10. Sensitivity analyses

#### 10A. Case-control numbers

**Supplement table 6.** Case-control numbers sensitivity analyses

| | | ≥ 2 bipolar disorder measures | | Bipolar disorder (MHQ) | |
|---|---|---|---|---|---|
| Dataset | Sex | Healthy control | Bipolar disorder | Healthy control | Bipolar disorder |
| Main dataset | Female | 132900 | 469 | 41131 | 693 |
| | Male | 135933 | 426 | 40563 | 488 |
| Lung function | Female | 46308 | 170 | 14586 | 245 |
| | Male | 54295 | 164 | 16779 | 193 |
| Bone mineral density | Female | 92461 | 268 | 26667 | 432 |
| | Male | 93165 | 235 | 26376 | 307 |
| Arterial stiffness | Female | 41238 | 212 | 14625 | 262 |
| | Male | 44301 | 201 | 14600 | 184 |

*Note:* MHQ = mental health questionnaire.

#### 10B. Case-control differences

**Supplement table 7.** Differences in physiological measures between individuals with bipolar disorder and healthy controls

| | **≥ 2 bipolar disorder measures** | | | | | **Bipolar disorder (MHQ)** | | | | |
|---|---|---|---|---|---|---|---|---|---|---|
| Variable | SMD | 95% CI | | $p_{Bonf.}$ | $p_{BH}$ | SMD | 95% CI | | $p_{Bonf.}$ | $p_{BH}$ |
| **Female** | | | | | | | | | | |
| Hand-grip strength | -0.216 | -0.306 | -0.125 | <0.001 | <0.001 | -0.066 | -0.141 | 0.009 | >0.999 | 0.182 |
| Systolic blood pressure | -0.196 | -0.287 | -0.105 | <0.001 | <0.001 | -0.263 | -0.338 | -0.188 | <0.001 | <0.001 |
| Diastolic blood pressure | -0.022 | -0.113 | 0.069 | >0.999 | 0.632 | -0.080 | -0.155 | -0.005 | 0.617 | 0.088 |
| Pulse rate | 0.173 | 0.083 | 0.264 | 0.010 | 0.001 | 0.073 | -0.003 | 0.148 | 0.996 | 0.124 |
| Body mass index | 0.372 | 0.281 | 0.462 | <0.001 | <0.001 | 0.325 | 0.250 | 0.400 | <0.001 | <0.001 |
| Body fat percentage | 0.281 | 0.191 | 0.372 | <0.001 | <0.001 | 0.202 | 0.127 | 0.277 | <0.001 | <0.001 |
| Fat mass | 0.340 | 0.249 | 0.431 | <0.001 | <0.001 | 0.285 | 0.210 | 0.360 | <0.001 | <0.001 |
| Fat-free mass | 0.273 | 0.182 | 0.364 | <0.001 | <0.001 | 0.254 | 0.179 | 0.330 | <0.001 | <0.001 |
| Waist-hip ratio | 0.430 | 0.340 | 0.521 | <0.001 | <0.001 | 0.171 | 0.096 | 0.246 | <0.001 | <0.001 |
| Peak expiratory flow | -0.039 | -0.189 | 0.112 | >0.999 | 0.632 | 0.062 | -0.064 | 0.189 | >0.999 | 0.378 |
| Forced expiratory volume 1s | -0.150 | -0.300 | 0.001 | 0.970 | 0.097 | 0.041 | -0.085 | 0.168 | >0.999 | 0.518 |
| Forced vital capacity | -0.152 | -0.303 | -0.001 | 0.574 | 0.064 | 0.016 | -0.111 | 0.142 | >0.999 | 0.795 |
| $FEV_1$ / FVC | -0.055 | -0.206 | 0.095 | >0.999 | 0.632 | 0.088 | -0.039 | 0.214 | >0.999 | 0.216 |
| Heel bone mineral density | -0.041 | -0.161 | 0.079 | >0.999 | 0.632 | 0.067 | -0.028 | 0.162 | >0.999 | 0.250 |
| Arterial stiffness | -0.029 | -0.164 | 0.106 | >0.999 | 0.632 | 0.092 | -0.030 | 0.214 | >0.999 | 0.183 |
| **Male** | | | | | | | | | | |
| Hand-grip strength | -0.289 | -0.384 | -0.194 | <0.001 | <0.001 | 0.009 | -0.080 | 0.098 | >0.999 | 0.980 |
| Systolic blood pressure | -0.167 | -0.262 | -0.072 | 0.005 | 0.001 | -0.249 | -0.338 | -0.160 | <0.001 | <0.001 |
| Diastolic blood pressure | -0.136 | -0.231 | -0.041 | 0.110 | 0.014 | -0.003 | -0.093 | 0.086 | >0.999 | 0.980 |
| Pulse rate | 0.190 | 0.095 | 0.286 | 0.006 | 0.001 | 0.159 | 0.070 | 0.248 | 0.015 | 0.003 |
| Body mass index | 0.187 | 0.092 | 0.282 | 0.012 | 0.002 | 0.281 | 0.192 | 0.370 | <0.001 | <0.001 |
| Body fat percentage | 0.172 | 0.077 | 0.267 | 0.021 | 0.003 | 0.135 | 0.045 | 0.224 | 0.114 | 0.016 |
| Fat mass | 0.226 | 0.131 | 0.321 | 0.002 | 0.001 | 0.227 | 0.137 | 0.316 | 0.001 | <0.001 |
| Fat-free mass | 0.128 | 0.033 | 0.223 | 0.242 | 0.027 | 0.231 | 0.142 | 0.320 | <0.001 | <0.001 |
| Waist-hip ratio | 0.295 | 0.200 | 0.390 | <0.001 | <0.001 | 0.171 | 0.082 | 0.260 | 0.010 | 0.002 |
| Peak expiratory flow | -0.149 | -0.303 | 0.004 | 0.752 | 0.068 | 0.062 | -0.080 | 0.204 | >0.999 | 0.575 |
| Forced expiratory volume 1s | -0.144 | -0.297 | 0.009 | >0.999 | 0.103 | 0.096 | -0.046 | 0.238 | >0.999 | 0.285 |
| Forced vital capacity | -0.185 | -0.338 | -0.031 | 0.279 | 0.028 | 0.058 | -0.084 | 0.200 | >0.999 | 0.575 |
| $FEV_1$ / FVC | 0.025 | -0.128 | 0.179 | >0.999 | 0.780 | 0.104 | -0.038 | 0.246 | >0.999 | 0.184 |
| Heel bone mineral density | -0.094 | -0.222 | 0.034 | >0.999 | 0.148 | 0.017 | -0.096 | 0.129 | >0.999 | 0.980 |
| Arterial stiffness | 0.039 | -0.100 | 0.177 | >0.999 | 0.488 | 0.002 | -0.144 | 0.147 | >0.999 | 0.980 |

*Note:* SMD = standardised mean difference; CI = confidence interval; Bonf. = Bonferroni; BH = Benjamini & Hochberg; $FEV_1$ = forced expiratory volume in one second; FVC = forced vital capacity; MHQ = mental health questionnaire. *P*-values for Welch's t-test.

10C. Age-related changes in females

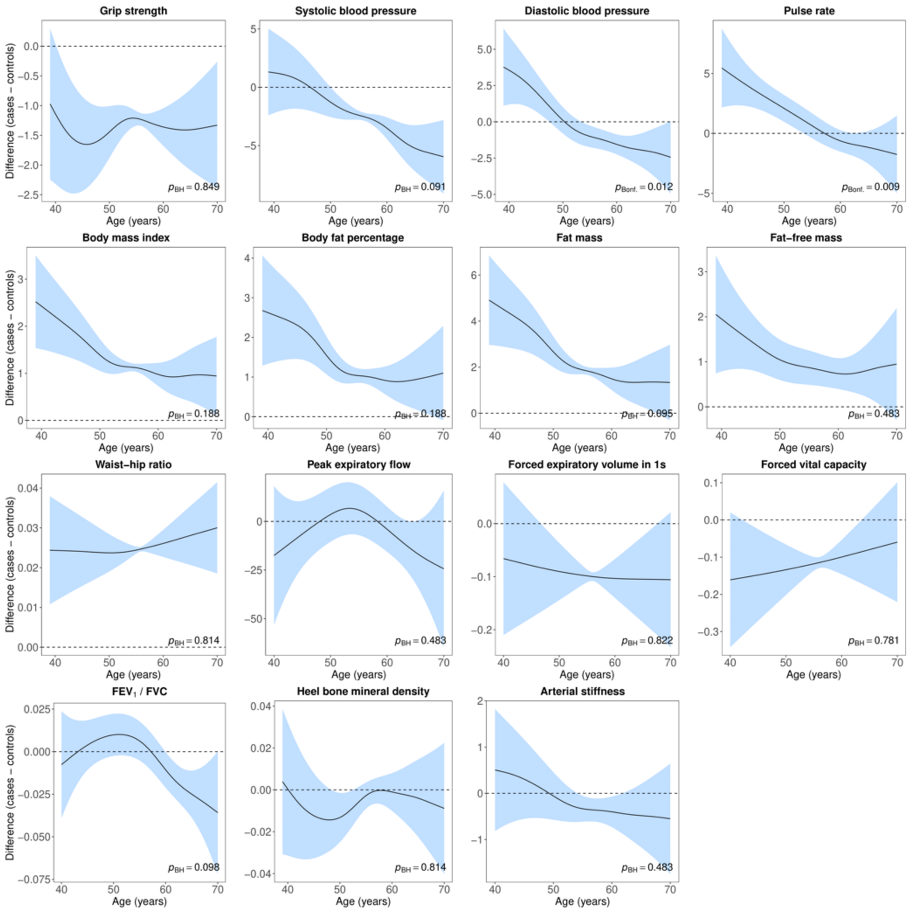

**Supplement figure 5.** Difference smooths comparing age-related changes in physiological measures of females with bipolar disorder (**≥ 2 bipolar disorder measures**) to healthy controls. Models were adjusted for ethnicity (except lung function), gross annual household income, physical activity, smoking status, alcohol intake frequency, sleep duration and, for cardiovascular measures, current use of antihypertensive medications. The shaded areas correspond to approximate 95% confidence intervals (± 2 × standard error). Negative values on the y-axes correspond to lower values in females with bipolar disorder compared to healthy controls. The horizontal lines represent no difference between female cases and controls. $FEV_1$ = forced expiratory volume in one second; FVC = forced vital capacity; Bonf. = Bonferroni; BH = Benjamini & Hochberg.

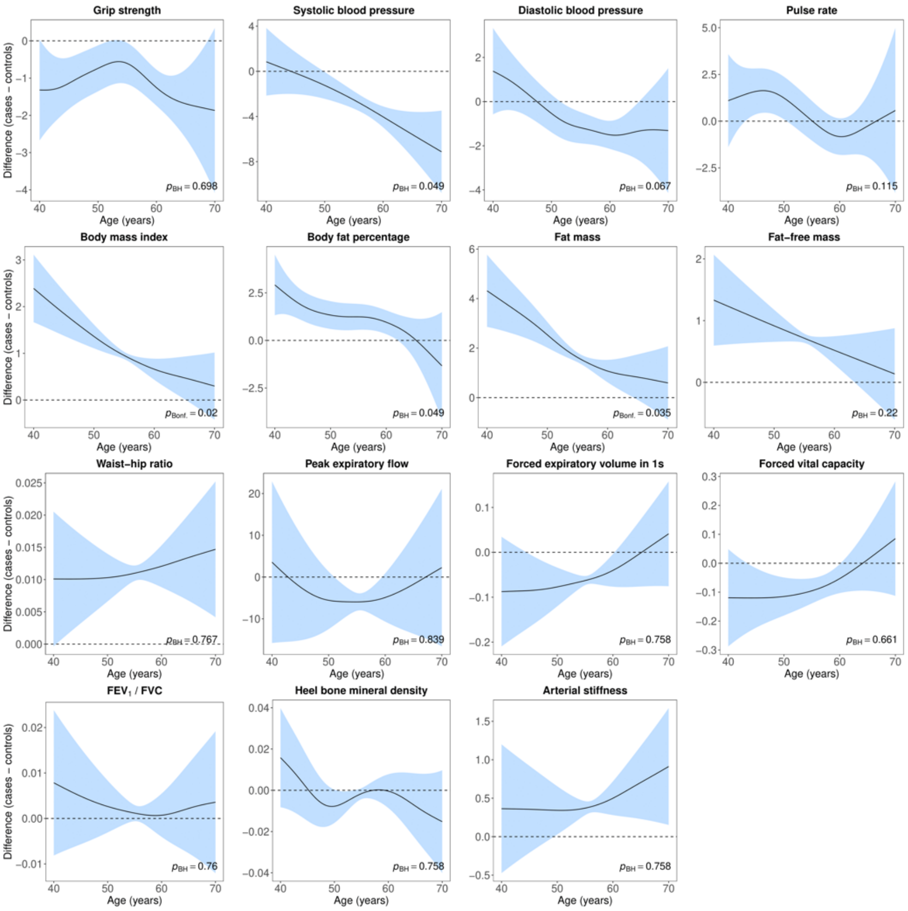

**Supplement figure 6.** Difference smooths comparing age-related changes in physiological measures of females with bipolar disorder (**MHQ**) to healthy controls. Models were adjusted for ethnicity (except lung function), gross annual household income, physical activity, smoking status, alcohol intake frequency, sleep duration and, for cardiovascular measures, current use of antihypertensive medications. The shaded areas correspond to approximate 95% confidence intervals (± 2 × standard error). Negative values on the y-axes correspond to lower values in females with bipolar disorder compared to healthy controls. The horizontal lines represent no difference between female cases and controls. $FEV_1$ = forced expiratory volume in one second; FVC = forced vital capacity; Bonf. = Bonferroni; BH = Benjamini & Hochberg.

10D. Age-related changes in males

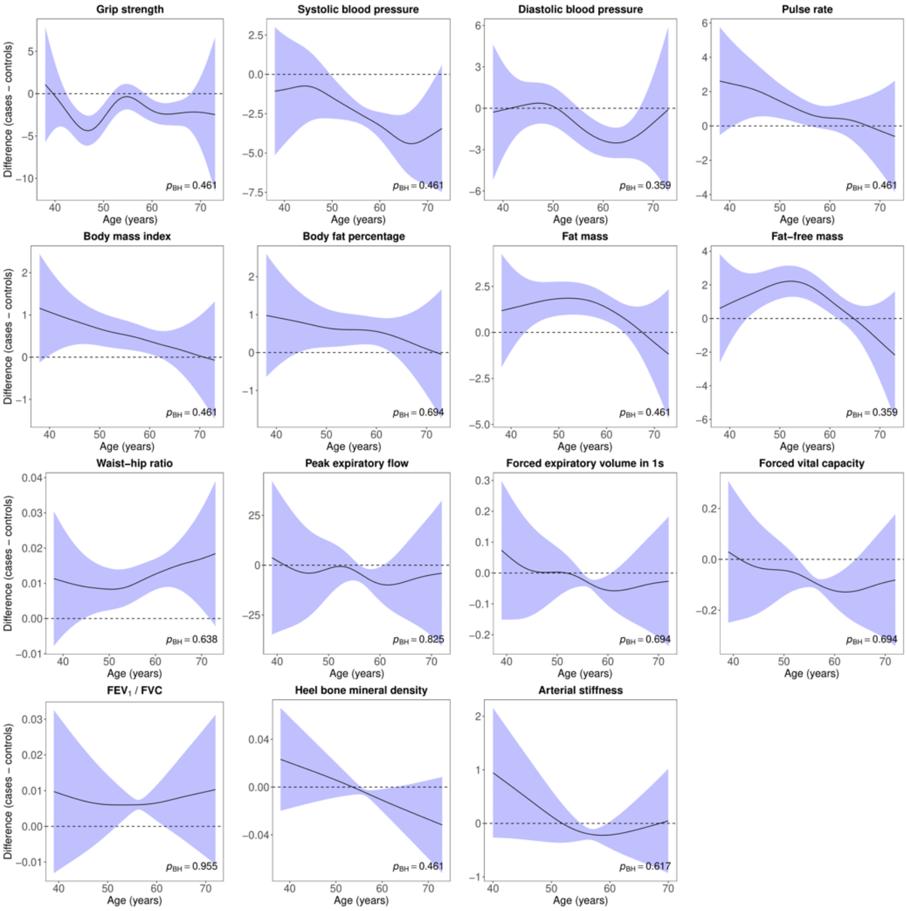

**Supplement figure 7.** Difference smooths comparing age-related changes in physiological measures of males with bipolar disorder (**≥ 2 bipolar disorder measures**) to healthy controls. Models were adjusted for ethnicity (except lung function), gross annual household income, physical activity, smoking status, alcohol intake frequency, sleep duration and, for cardiovascular measures, current use of antihypertensive medications. The shaded areas correspond to approximate 95% confidence intervals (± 2 × standard error). Negative values on the y-axes correspond to lower values in males with bipolar disorder compared to healthy controls. The horizontal lines represent no difference between male cases and controls. $FEV_1$ = forced expiratory volume in one second; FVC = forced vital capacity; Bonf. = Bonferroni; BH = Benjamini & Hochberg.

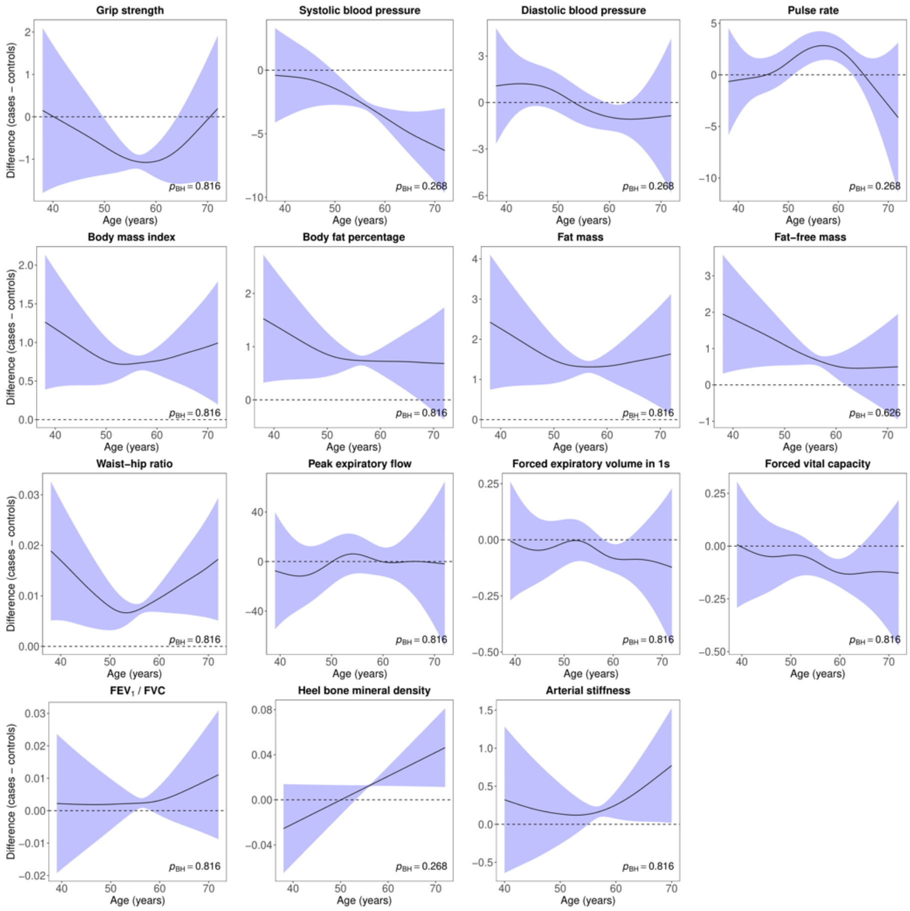

**Supplement figure 8.** Difference smooths comparing age-related changes in physiological measures of males with bipolar disorder (**MHQ**) to healthy controls. Models were adjusted for ethnicity (except lung function), gross annual household income, physical activity, smoking status, alcohol intake frequency, sleep duration and, for cardiovascular measures, current use of antihypertensive medications. The shaded areas correspond to approximate 95% confidence intervals (± 2 × standard error). Negative values on the y-axes correspond to lower values in males with bipolar disorder compared to healthy controls. The horizontal lines represent no difference between male cases and controls. $FEV_1$ = forced expiratory volume in one second; FVC = forced vital capacity; Bonf. = Bonferroni; BH = Benjamini & Hochberg.
